# Characterization and forecast of global influenza (sub)type dynamics

**DOI:** 10.1101/2024.08.01.24311336

**Authors:** Francesco Bonacina, Pierre-Yves Boëlle, Vittoria Colizza, Olivier Lopez, Maud Thomas, Chiara Poletto

## Abstract

The (sub)type composition of seasonal influenza waves varies in space and time. (Sub)types tend to have different impacts on population groups; therefore, understanding the drivers of their co-circulation and anticipating their composition is important for epidemic preparedness. FluNet provides data on influenza specimens by (sub)type for more than 150 countries. However, due to surveillance variations across countries, global analyses usually focus on (sub)type compositions, a kind of data difficult to treat with advanced statistical methods. We used Compositional Data Analysis to circumvent the problem and study trajectories of annual (sub)type compositions of countries. First, we examined global trends from 2000 to 2023. We identified a few seasons which stood out for the strong within-country (sub)type dominance due to either a new virus/clade taking over (2003/2004 season, A/H1N1pdm pandemic) or (sub)types’ spatial segregation (COVID-19 pandemic). Second, we showed that geographical factors, most notably international mobility, concurred in shaping countries’ composition trajectories between 2010 and 2019. Trajectories clustered in two macroregions characterized by (sub)type alternation vs. persistent mixing. Finally, we defined five algorithms for forecasting the next year’s composition and found that incorporating the global history of (sub)type composition in a Bayesian Hierarchical Vector AutoRegressive model improved predictions compared with naive methods. The joint analysis of spatiotemporal dynamics of influenza (sub)types worldwide revealed a hidden structure in (sub)type circulation that can be used to improve predictions of the (sub)type composition of next year’s epidemic according to place.

## Introduction

Since 2009, the H1N1pdm and H3N2 subtypes of influenza type A and influenza type B have been co-circulated in the human population, with highly variable occurrences in space and time (1, 2). The viral diversity profoundly impacts the epidemiological characteristics of epidemics: epidemics dominated by A/H3N2 strains are generally more severe as this strain is more transmissible (3) and causes increased morbidity in the elderly due to the immune imprinting mechanism (4–6); while epidemics due to the A/H1N1 and B strains tend to cause more infections in the young (6–8). The diversity of strains also manifests with extended epidemic periods, as, for example, the peak of A infections and of B infections are typically separated by a few weeks in the northern temperate regions (9, 10). Anticipating the (sub)type composition of the coming season is therefore key to improving preparedness, e.g., planning hospital capacities, awareness campaigns, and vaccine distribution (10, 11).

Yet, anticipating the (sub)type co-circulation is complicated because influenza viruses interact with one another. Evidence of viral interference has been found experimentally (3, 12), from mathematical models fitted to country-level incidence data (13, 14), and from country (10, 11) and multi-country (9, 15, 16) statistical analyses. In particular, past studies have shown that cross-immunity is an important ingredient of models aiming at reproducing plausible influenza dynamics (13, 14). In other words, Influenza (sub)types form a coupled ecological system that needs to be studied as a whole. A second source of complication is represented by the fact that influenza rapidly spreads globally (17, 18) and epidemics in different countries are interdependent. This makes the study of worldwide influenza circulation critical for interpreting the viral patterns observed within a focal country.

In response to these needs, the Global Influenza Surveillance and Response System (GISRS) (1) gathers and makes available through the FluNet portal the weekly number of samples by (sub)types and country. The quality and quantity of the data are constantly increasing, but (sub)typing effort is not standardized across countries yet. To allow multi-countries comparison, it is therefore preferable to focus on rescaled quantities, such as the percentages of infections by each (sub)type. These data allowed for showing strong patterns of alternation between A/H1N1pdm and A/H3N2 in Europe (19, 20), the domination of A/H3N2 among all (sub)types and inter-hemispheric synchrony in its circulation (9). It was also used to characterize the altered (sub)type circulation following the emergence of the A/H1N1pdm (sub)type (15) and the COVID-19 pandemic (21). However, these studies did not address the spatial drivers of (sub)type composition nor provide anticipation. Research work attempting to forecast the next year’s (sub)type composition in a country is so far scarce (22).

Here, we undertook a systematic analysis of the coupled dynamics of (sub)types by analyzing their relative abundances across countries and years through the Compositional Data Analysis (CoDA) framework (23, 24). This approach has been used in ecology and geology to analyze percentage data, taking into account the sum-to-one constraint. Yet, its use in infectious disease dynamics has been notably rare. Here, we defined a trajectory in the (sub)type composition space for each country. We quantified how these trajectories evolved in time and space and identified predictors of spatial structure. We then proposed an approach to leverage this structure to forecast (sub)type relative abundance in each country one year ahead with improved accuracy compared with naive estimators.

## Results

### Studying the relative abundances of influenza (sub)types with Compositional Data Analysis (CoDA)

We studied the relative abundance of influenza (sub)types for different countries/years, defined by the percentages of the form (B%, H1%, H3%). For brevity, we will use in figures and equations H1 for the A/H1N1 strains - historical A/H1N1 before 2009, and the A/H1N1pdm after -, and H3 for A/H3N2. We consider weekly surveillance data reported in FluNet (1, 25) from 2000 to 2024, for up to 167 countries. We aggregated data annually, from May to April, and computed “compositions” (23), i.e., the percentages of each of the three (sub)types for each year and country (Fig. 1A) (for details see the Methods). These compositions are “sum to one” data that can be shown in a ternary plot (Fig. 1B).

**Fig. 1.**
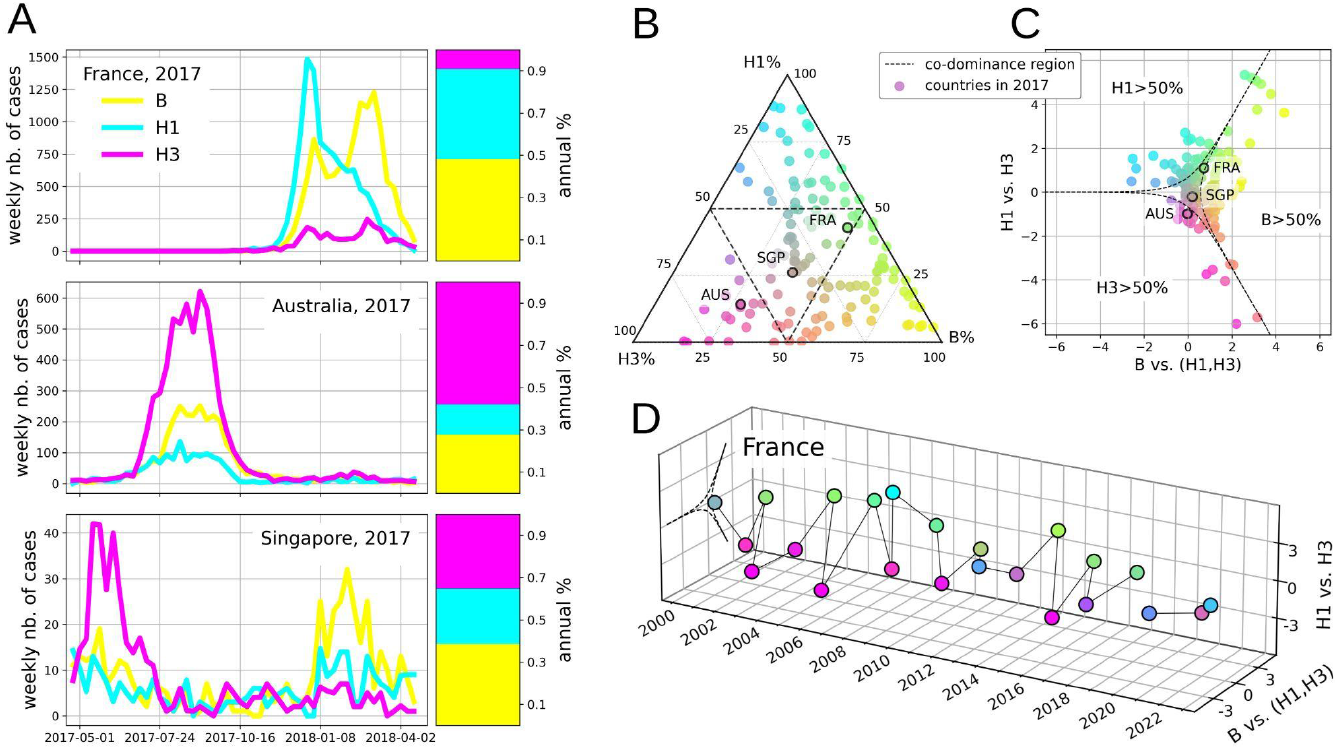
Use of CoDA to define (sub)type abundance trajectories. A) FluNet samples by (sub)types for France, Australia, and Singapore. We consider the year 2017, defined as the period going from May 2017 to Apr 2018, as an example and compute the (sub)type abundances over the year. B) Proportions of A/H1N1 strains (A/H1N1 before 2009, and A/H1N1pdm after 2009), A/H3N2, and B represented in a ternary graph. Coloured points represent the observations for each of the 138 countries considered in 2017. The dotted line separates the three dominance and the (central) co-dominance regions. Points are colour-coded according to the composition in B (yellow), A/H1N1 (cyan), and A/H3N2 (magenta) - grey indicates a perfect balance of circulating strains. C) Relative abundances of (sub)types of the same 138 points plotted in panel B) after isometric log-ratio transformation. D) Trajectory of relative abundances log-ratios in France, from 2000 to 2023. The point for 2020 is missing because fewer than 50 cases were uploaded on FluNet that year. Points are coloured with the same triangular code as in Fig. B) and C). In the figure, H3 is used to indicate A/H3N2, and H1 is used to indicate A/H1N1 before 2009, and A/H1N1pdm after 2009.

The study of this type of data is complicated since the components are inherently non-independent (23, 26), making standard statistical tools inadequate (24). Modern compositional analysis solves the issue by transforming the original data to log-ratios (27). Here, we used the isometric log-ratio transformation (*ilr*) (27). An alternative, the additive log-ratio transformation, was also tested in the Supplementary Information. The *ilr* transformation is as follows:

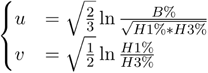

While analyses performed after CoDA transformation are invariant under any permutation of percentages, we chose to use the log-ratio of B% to 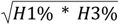 and of H1% to H3%, as it provides a straightforward epidemiological interpretation. The ternary graph can then be remapped to these new coordinates (Fig. 1C), which makes it easy to represent trajectories of (sub)type abundance over time for a given country (Fig. 1D). In these graphs, we also highlighted “dominance” boundaries corresponding to compositions with one (sub)type amounting to more than 50% of the samples. The central region, or “co-dominance” region, corresponds to neither (sub)type being dominant.

### Global multiannual trends in (sub)type mixing

The composition trajectories for 167 countries are shown in Fig. 2A. The number of countries contributing to FluNet was initially limited, but has substantially increased since then. For example, in 2000, only 31 countries contributed more than 50 samples, our threshold for inclusion, compared to 149 countries in 2023.

**Fig. 2.**
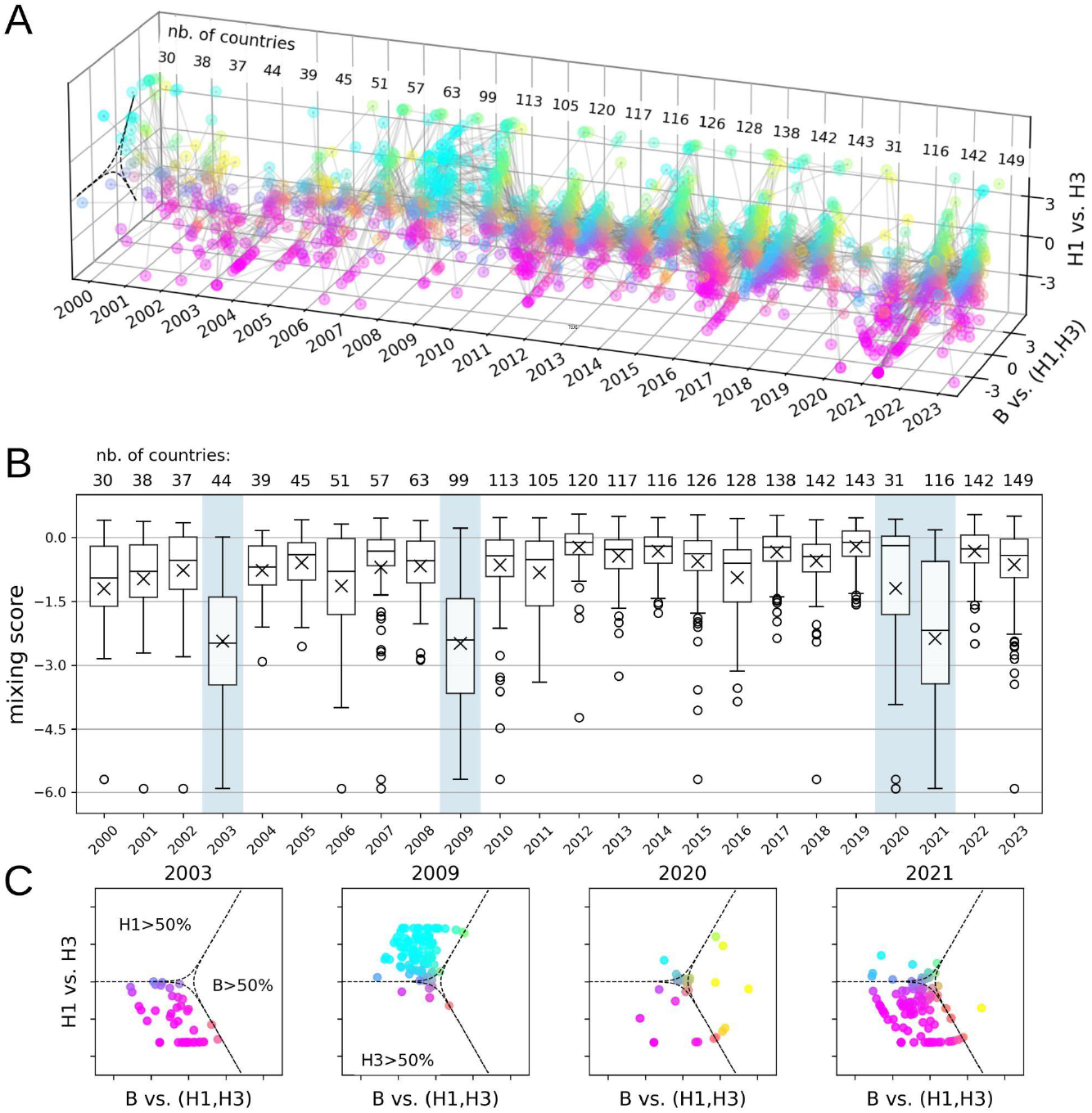
Trajectories of relative abundances of influenza (sub)types and global (sub)type mixing. A) Trajectories of relative abundances of influenza (sub)types. We present trajectories for 167 countries, including years from 2000 to 2023. Points in the plot represent countries/years for which the number of FluNet cases is at least 50. The number of countries included by year is shown at the top. B) Degree of mixing of influenza (sub)types over time. For the years 2000 to 2023, the mixing score of influenza (sub)types was computed for each country, and their distributions are depicted through the boxplots. Positive scores represent countries where each (sub)type is responsible for <50% of cases, while negative scores denote the dominance of one (sub)type. C) Influenza (sub)type abundances for 2003, 2009, 2020, and 2021, when (sub)type mixing was unusually low. In the figure, H3 is used to indicate A/H3N2, and H1 is used to indicate A/H1N1 before 2009, and A/H1N1pdm after 2009. Points are colored with the same triangular code as in Fig. 1.

We first analyzed global statistics on the ensemble of trajectories from one year to another. We introduced the *mixing score* to quantify the (sub)type mixing in a country/year. This is defined as the shortest distance in the log-ratio plane between the country/year composition and the boundary of the co-dominance region. It ranges from positive values when the composition is inside the co-dominance region to negative ones when it is outside, i.e., one (sub)type is dominant. The measure has an upper bound at 0.56, corresponding to the equipartition of cases of a country/year among the three (sub)types, and no lower bound. For example, strong dominance—e.g., one (sub)type exceeding 75% frequency—yields a mixing score below, e.g., -0.8, depending on how the remaining two (sub)types are distributed. The distribution of countries’ mixing scores in a given year provides a global overview of country-level (sub)type mixing. The multiannual comparison of Fig. 2B highlights anomalous events, when the mixing score was extraordinarily low, overall. A/H3N2 was strongly dominant (i.e. >75%) in 35 out of 44 countries in 2003, and A/H1N1pdm was strongly dominant in 82 out of 99 countries in 2009 (Fig. 2C). One (sub)type among A/H1N1pdm, A/H3N2, and B was strongly dominant in 13 out of 31 countries in 2020, and in 83 out of 116 countries in 2021.

Atypical values correspond to events occurring at the global scale in those years. The strong dominance of A/H3N2 in 2003 was caused by an emerging clade for which the vaccine had limited effect (28, 29). Similarly, the strong dominance of A/H1N1pdm in 2009 was due to the zoonotic emergence of the A/H1N1pdm subtype and the consequent pandemic (15, 30). In both examples, a punctuated change in the virus caused a strong level of dominance, which was associated with a high level of subtype synchrony across countries, and more intense viral circulation globally. During 2020–2021, (sub)types circulated with enhanced regional segregation (31). In 2020, A/H3N2 strongly dominated in seven Southeast Asian countries, B in five tropical-belt countries, and A/H1N1pdm circulated mainly in five African countries, Mexico, and South Korea (Supplementary Information, Fig. S3). This was due to the unprecedented contact and mobility restrictions during COVID-19 (31–33), which caused a worldwide drop in influenza cases (only 31 countries reported at least 50 samples in 2020). In the Supplementary Information, we analyzed the mixing score under additive log-ratio coordinates. We also show that the Shannon entropy provides a result similar to the mixing score. The advantage of the mixing score is that it provides an easy-to-read visualization of how far the composition is from co-dominance.

### Geographical patterns in (sub)type cocirculation

We then looked for geographical patterns in countries’ trajectories from 2010 to 2019, the longest recent period of regular influenza circulation, between the 2009 A/H1N1pdm and the COVID-19 pandemics, and characterized by a large number of countries (86) satisfying our inclusion criteria. To identify predictors of country compositions, we took distances between countries’ trajectories in the log-ratio plane as the response variable and applied both univariable correlations and multivariable linear regression. Predictors included country-pair differences in average temperature and humidity, inter-country air connectivity, and epidemic synchrony—classified as synchronous (same seasonal peak), asynchronous (opposite hemispheres), or semi-synchronous (seasonal vs. year-round circulation). See Methods and Supplementary Information for details. All covariates were statistically significant (Fig. 3A, Supplementary Information, Tables S2). Among the three continuous covariates, air traffic had the strongest effect. Synchrony was also very important.

**Fig. 3.**
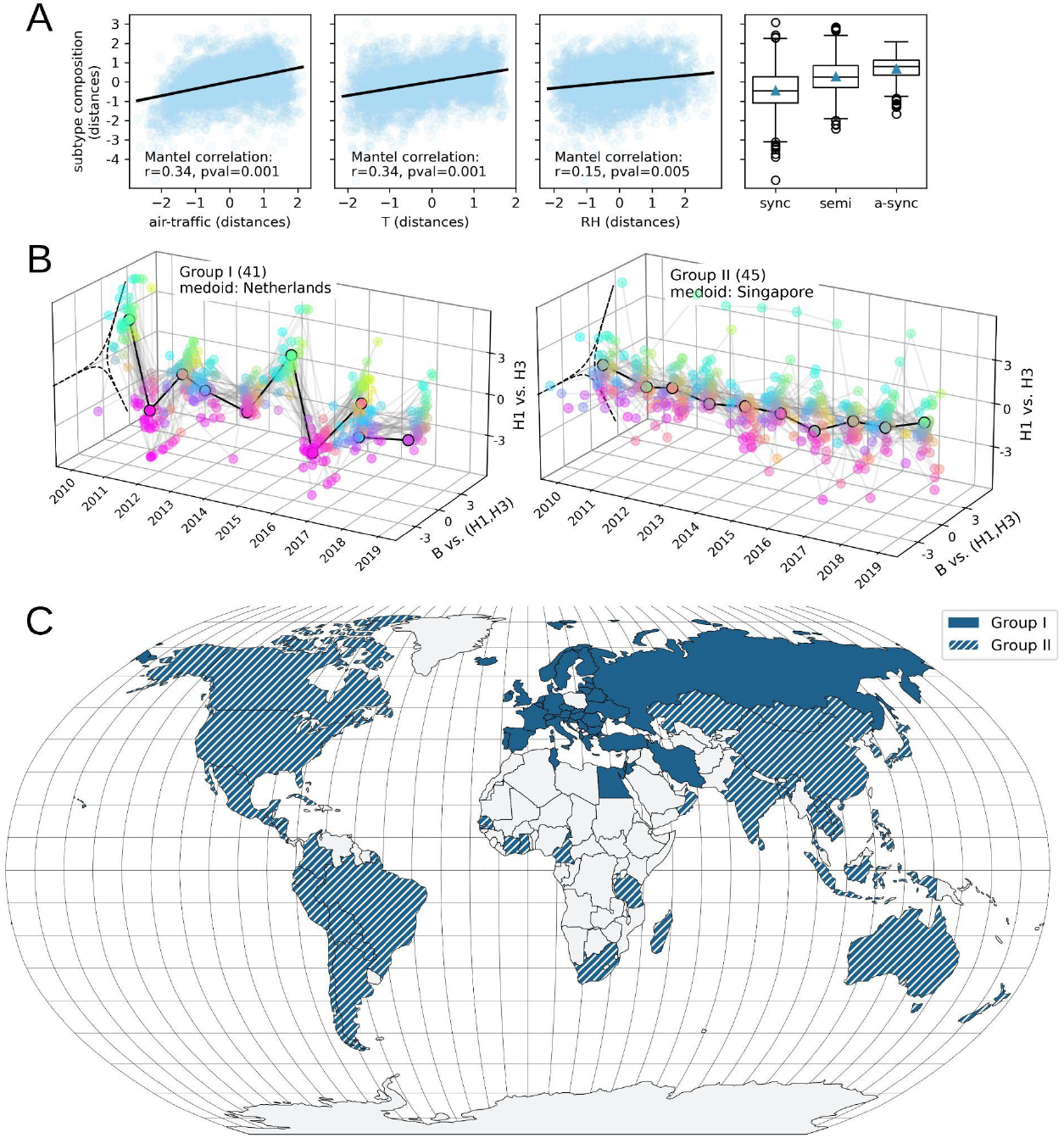
Geographical pattern of (sub)type trajectories. A) Association of country-pair (sub)type trajectory distances with air traffic, temperature, relative humidity, and epidemic synchrony. Continuous variables were standardized, and their correlation with the response variable was assessed using the Mantel test, with results reported in the legends. Linear models are also shown to highlight trends. B) Hierarchical clustering of trajectories of A/H1N1pdm, A/H3N2, and B relative abundances. Trajectories for 86 countries from 2010 to 2019 are clustered in two groups - groups I and II - with 41 and 45 countries, respectively. The medoid trajectories - i.e., the most central trajectories - of the two groups are highlighted in black and are reported in the legends. The co-dominance regions are shown with dashed lines as a reference. C) Geographic positioning of Group I and Group II countries. Source of shape files for the map: Natural Earth (35). In the figure, H3 is used to indicate A/H3N2, and H1 is used to indicate A/H1N1pdm. Points are coloured with the same triangular code as in Fig. 1.

To visualize the global pattern of countries’ compositions, we clustered countries with similar trajectories. Fig. 3B shows the clustering in two groups. These groups comprised whole WHO Influenza Transmission Zones (ITZs), with only 3 countries allocated to the other group (34). Group I included 41 countries from the WHO ITZs of Europe, Northern Africa, and Western Asia (Supplementary Information, Fig. S4), whose trajectories showed strong and synchronous alternation between (sub)types (Fig. 3B). The 45 countries of Group II belonged to the other WHO ITZs, except for Oman and Qatar from the Western Asian ITZ (Supplementary Information, Fig. S4). These countries displayed flatter trajectories overall, though with more variability between them than seen in Group I. (Fig. 3B). Group II included tropical countries characterized by co-circulation of all (sub)types throughout the year, seven northern temperate countries exhibiting marked (sub)type alternation, yet not synchronous with Group I (the USA, Canada, China, Hong Kong SAR of China, Mongolia, Republic of Korea, Japan), and countries from South and Central America (Supplementary Information, Fig. S4). A finer clustering in six groups split Group I into two (roughly Western Asia vs. Europe) and Group II into the three previously described subgroups, plus Qatar (Supplementary Information, Figs. S4 and S5), here again in good accordance with the WHO ITZs. For both the two-groups and the six-groups repartition, countries belonging to the same cluster showed more similar temperatures and/or increased connection and/or synchrony compared to the whole set of countries (Supplementary Information, Fig. S6).

The analysis of predictors of geographical patterns was robust under changes in the log-ratio transformation. When an alternative influenza year beginning in September was tested, results were similar except that the difference between the trajectories of asynchronous countries was smaller than in the baseline. Under a stricter criterion for inclusion of countries/years, we found that temperature and relative humidity were no longer statistically significant, whereas the influence of air travel increased (Supplementary Information, Table S4). This may be due to the exclusion of many countries in this scenario - mainly those in the tropical belt - for which similarities in (sub)type composition were better explained by climate than by air connections (Supplementary Information, Fig. S6). The country grouping was robust in all sensitivity scenarios tested (Supplementary Information).

### Forecasting of next year’s influenza (sub)type composition

By examining (sub)type compositions as trajectories over time, we can try to predict next year’s influenza (sub)type composition or dominance pattern. As there is currently no accepted approach to carry out this task, we considered several possibilities based on epidemiological considerations or using state-of-the-art statistical models building on the CoDA framework. We focused on predicting compositions for 2017, 2018, and 2019 using the history of compositions observed worldwide since 2010. The models were as follows (see Methods for details):

- *M1 past frequencies*: prediction for the coming year corresponds to the dominance/co-dominance state - between dominance of A/H1N1pdm, A/H3N2, B, or co-dominance - that occurred most often in previous years. Note that it does not predict a composition but only the dominance/co-dominance state.
- *M2 H1-H3 alternation:* we use the composition of the past year in the same country, exchanging the percentages of A/H1N1pdm and A/H3N2. This is based on the evidence that A/H1N1pdm and A/H3N2 viruses tend to alternate, while influenza B often co-dominates (9, 15, 20).
- *M3 average composition:* we compute the average of previous years’ compositions in the same country, after the log-ratio transformation.
- *M4 VAR*: we fit a Vector AutoRegressive (VAR) model, which predicts the future composition of a country as a linear combination of its past compositions, accounting for both temporal autocorrelation and correlations between components (36). We used a VAR model with lag=1.
- *M5 HVAR*: we fit a Bayesian Hierarchical Vector AutoRegressive (HVAR) model to provide predictions for each individual country from past compositions of countries with similar trajectories (37). The approach extends the *M4 VAR* model and compensates for limited data over time by incorporating information on (sub)type compositions over space. In practice, we first clustered the training set trajectories as described in the previous section. Then, an HVAR model was fitted separately to each cluster, assuming that while each trajectory follows a VAR process, the VAR parameters within a cluster are not too dissimilar. We tested both lag=1 and lag=2. Fitted VAR coefficients are analysed in the Supplementary Information.

From models *M3, M4*, and *M5*, it was also possible to compute prediction regions based on 95% probability intervals. Models *M1* and *M2* only allowed point predictions.

Table 1, top panel, shows the forecast performances of the models by looking at the percentages of correctly predicted dominance states - the *Dominance State Accuracy*. Results are presented globally and for Group I and Group II countries. Model *M1* had 27% accuracy overall, performing similarly to random guessing the dominance state out of the four possibilities, i.e., dominance of A/H1N1pdm, A/H3N2, B, or co-dominance. Accuracy was lower for Group I (24%) than for Group II (30%), consistent with the difficulty in predicting the marked (sub)type alternation of Group I countries. Other models (*M2, M3*, and *M4*) had performances close to model *M1*. Estimates improved with *M5 HVAR*, with a global accuracy of 34%, improving in both Group I (28%) and Group II (40%).

**Table 1.** Evaluating influenza (sub)type forecasting. Scores considered to compare the five predictive models are the *Dominance State Accuracy* for dominance state predictions and the *Energy Score* for predictions of compositions. Scores are averaged over predicted years (2017, 2018, 2019) and countries within the same group - i.e., Group I, Group II, and all the countries, are reported in different rows. Scores are reported with uncertainties expressed as Standard Errors of the Mean (SEM) calculated over 200 bootstrap samples. For the *M5 HVAR* model, we report the result with the lag that performed the best for each group, i.e., lag=2 for Group I and lag=1 for Group II. Methods that performed best by country groupings are in bold for both the *Dominance State Accuracy* and the *Energy Score. Dominance State Accuracy* is positively oriented (i.e., it increases as the model’s performance increases), whereas the *Energy Score* is negatively oriented (i.e., it decreases as the model’s performance increases).

| Model | M1 past frequencies | M2 H1-H3 alternation | M3 average composition | M4 VAR | M5 HVAR |
| --- | --- | --- | --- | --- | --- |
| **Predicted observable: dominance state** |  |  |  |  |  |
| **Score: Dominance State Accuracy** |  |  |  |  |  |
| **Group I** | 0.235 ± 0.003 | 0.227 ± 0.003 | 0.218 ± 0.003 | 0.193 ± 0.003 | **0.277 ± 0.003** |
| **Group II** | 0.301 ± 0.003 | 0.301 ± 0.003 | 0.353 ± 0.003 | 0.257 ± 0.003 | **0.397 ± 0.003** |
| **All countries** | 0.271 ± 0.002 | 0.267 ± 0.002 | 0.290 ± 0.002 | 0.227 ± 0.002 | **0.341 ± 0.002** |
| **Predicted observable: composition** |  |  |  |  |  |
| **Score: Energy Score** |  |  |  |  |  |
| **Group I** | / | 3.813 ± 0.010 | 1.811 ± 0.008 | 2.194 ± 0.007 | **1.426 ± 0.005** |
| **Group II** | / | 2.102 ± 0.009 | 1.338 ± 0.007 | 1.640 ± 0.009 | **1.195 ± 0.005** |
| **All countries** | / | 2.901 ± 0.008 | 1.559 ± 0.006 | 1.899 ± 0.006 | **1.303 ± 0.004** |

Models *M2, M3, M4*, and *M5*, which generate continuous predictions of (sub)type compositions, were also compared using probabilistic forecasting metrics commonly adopted in epidemiology (38), where both calibration (closeness to the target) and sharpness (prediction uncertainty) are evaluated. In Table 1, bottom panel, we show the values of the *Energy Score* (39), where smaller values correspond to better predictions. We found again that the *M5 HVAR* model performed better than other models. Predictions for France (Group I) and Australia (Group II) using this model are illustrated in Fig. 4 as examples.

**Fig. 4.**
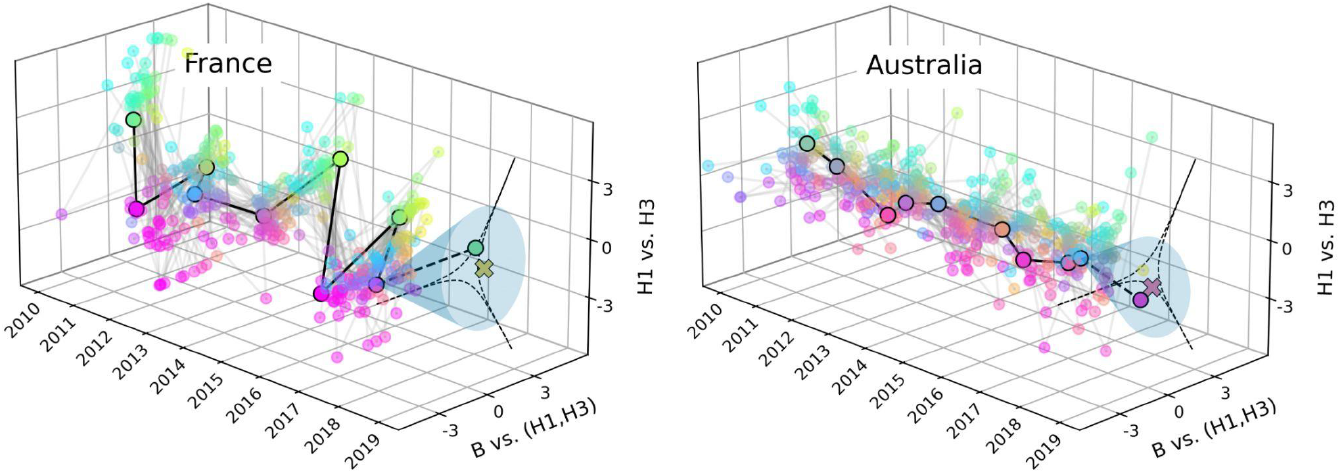
Prediction of relative abundances of influenza (sub)types for France and Australia in 2019. Predictions are computed using the *M5 HVAR* model of lag 2 and 1 (see Methods), respectively. For each country, the observed trajectory from 2010 to 2018 is represented with a black solid line, while the dashed segment links the points corresponding to 2019, to be predicted, to the rest of the time series. The thinner grey lines correspond to the trajectories of the other countries within the respective group - Group I for France, and Group II for Australia - that were used to train the model. The crosses depict the predictions and the shadow areas of the ellipses associated with the 95% confidence intervals. In the figure, H3 is used to indicate A/H3N2, and H1 is used to indicate A/H1N1pdm. Points are coloured with the same triangular code as in Fig. 1. The co-dominance regions are shown with dashed lines as a reference. The *Energy Scores* for the 2019 predictions for France and Australia are 0.90 and 0.66, respectively.

Finally, we addressed simpler prediction tasks still relevant for public health. Precisely, for each (sub)type, we asked whether it would be (i) dominant (>50% of cases) or not, and (ii) negligible (<10% of cases) or not. For model *M1 past frequencies*, the probability of negligibility of a (sub)type corresponded to the proportion of times the (sub)type was negligible in the previous years. To evaluate the performance of the five models in carrying out the two tasks, we used the *Area Under the Receiver Operating Characteristic Curve (AUROC)*. Results are provided in Table 2. The *M5 HVAR* model was the best-performing model in predicting the negligibility of influenza type B, the dominance of subtype A/H1N1pdm, and both the dominance and negligibility of subtype A/H3N2. In particular, the *M5 HVAR* provided the best performance and the greatest improvement when applied to the trajectories of Group I countries, in contrast with the forecasts of dominance and composition being less good for this group, as discussed above. For these countries, the *AUROC* score for the *M5 HVAR* model reached 0.91 in predicting B negligibility, compared to scores of 0.35 to 0.61 obtained with other models. Similarly, it reached 0.76 (0.80) when predicting the dominance (negligibility) of A/H3N2, compared to 0.43-0.64 (0.38-0.62) for the other models. *M5 HVAR* was not always the best-performing model, notably, it was not always able to improve the predictions of the *M1 past frequencies* and *M3 average composition* models in predicting the dominance of B and the negligibility of A/H1N1pdm. For the dominance of A/H1N1pdm for Group I countries, no model performed significantly better than a random guess.

**Table 2.** Forecast evaluation for predicting the dominance (yes/no) and negligibility (yes/no) of (sub)types. Results are summarized in six panels: the three top ones focus on dominance and the three bottom ones on negligibility. As a score of prediction, we used the *Area Under the Receiver Operating Characteristic Curve (AUROC)*. Scores were averaged over predicted years (2017, 2018, 2019) and countries within the same group - i.e. Group I, Group II, and all the countries, are reported in different rows. Scores are reported with uncertainties expressed as Standard Errors of the Mean (SEM) calculated over 200 bootstrap samples. The probability of dominance and negligibility of (sub)types cannot be calculated with the *M2 H1-H3 alternation* model; therefore, the AUROC cannot be computed for this model. We highlight the method performing best by country grouping.

| Model | M1 past frequencies | M2 H1-H3 alternation | M3 average composition | M4 VAR | M5 HVAR |
| --- | --- | --- | --- | --- | --- |
| Predicted observable: B will be dominant (>50%) in the next season: yes/no? |  |  |  |  |  |
| Score: AUROC |  |  |  |  |  |
| Group I | 0.452 ± 0.004 | / | 0.495 ± 0.004 | 0.425 ± 0.005 | **0.641 ± 0.004** |
| Group II | **0.715 ± 0.006** | / | 0.693 ± 0.005 | 0.463 ± 0.008 | 0.368 ± 0.009 |
| All countries | 0.551 ± 0.003 | / | **0.624 ± 0.003** | 0.496 ± 0.003 | 0.618 ± 0.004 |
| Predicted observable: H1 will be dominant (>50%) in the next season: yes/no? |  |  |  |  |  |
| Score: AUROC |  |  |  |  |  |
| Group I | **0.489 ± 0.004** | / | 0.414 ± 0.004 | 0.327 ± 0.004 | 0.480 ± 0.004 |
| Group II | 0.440 ± 0.004 | / | 0.505 ± 0.004 | 0.527 ± 0.004 | **0.584 ± 0.004** |
| All countries | 0.481 ± 0.003 | / | 0.485 ± 0.003 | 0.439 ± 0.003 | **0.529 ± 0.003** |
| **Predicted observable: H3 will be dominant (>50%) in the next season: yes/no?** |  |  |  |  |  |
| **Score: AUROC** |  |  |  |  |  |
| Group I | 0.433 ± 0.004 | / | 0.478 ± 0.005 | 0.640 ± 0.004 | **0.764 ± 0.003** |
| Group II | 0.552 ± 0.004 | / | 0.576 ± 0.004 | 0.511 ± 0.004 | **0.618 ± 0.004** |
| All countries | 0.506 ± 0.003 | / | 0.539 ± 0.003 | 0.578 ± 0.003 | **0.702 ± 0.003** |
| **Predicted observable: B will be negligible (<10%) in the next season: yes/no?** |  |  |  |  |  |
| **Score: AUROC** |  |  |  |  |  |
| Group I | 0.412 ± 0.004 | / | 0.349 ± 0.003 | 0.610 ± 0.004 | **0.905 ± 0.002** |
| Group II | 0.521 ± 0.006 | / | 0.507 ± 0.005 | 0.527 ± 0.004 | **0.648 ± 0.004** |
| All countries | 0.561 ± 0.003 | / | 0.506 ± 0.003 | 0.603 ± 0.003 | **0.815 ± 0.002** |
| **Predicted observable: H1 will be negligible (<10%) in the next season: yes/no?** |  |  |  |  |  |
| **Score: AUROC** |  |  |  |  |  |
| Group I | **0.699 ± 0.005** | / | 0.645 ± 0.006 | 0.281 ± 0.006 | 0.253 ± 0.006 |
| Group II | 0.588 ± 0.004 | / | 0.580 ± 0.004 | 0.548 ± 0.004 | **0.605 ± 0.006** |
| All countries | **0.596 ± 0.003** | / | 0.567 ± 0.004 | 0.423 ± 0.004 | 0.460 ± 0.004 |
| **Predicted observable: H3 will be negligible (<10%) in the next season: yes/no?** |  |  |  |  |  |
| **Score: AUROC** |  |  |  |  |  |
| Group I | 0.375 ± 0.004 | / | 0.447 ± 0.005 | 0.715 ± 0.004 | **0.796 ± 0.003** |
| Group II | 0.620 ± 0.004 | / | 0.553 ± 0.005 | 0.654 ± 0.006 | **0.655 ± 0.005** |
| All countries | 0.565 ± 0.003 | / | 0.587 ± 0.003 | 0.712 ± 0.003 | **0.758 ± 0.003** |

Results on model ranking were overall consistent when evaluating models with alternative metrics and when performing predictions under alternative scenarios - i.e., using alternative log-ratio coordinates, a different definition of the influenza year, restricting to countries/years with at least 500 cases, and the 50% of countries that were best classified into the two clusters (see Supplementary Information, Tables S1, S2, S5-S7). The forecast for the 50% best-classified countries outperformed the baseline for seven out of eight observables. Notably, AUROC improved from 0.62 to 0.82 in predicting the dominance of B with M5 HVAR, the best-performing model in this scenario. Stronger similarity with other countries in the same cluster may have improved the quality of the spatial information used for the fit in the HVAR model. Predictions for all countries and years, obtained with the five methods, are reported in the SI Dataset S1.

## Discussion

Viral composition plays a key role in shaping influenza dynamics and the population burden of epidemics. A better anticipation of seasonal epidemics requires an improved ability to monitor, quantify, and predict (sub)type composition. This is increasingly within our reach thanks to the rapid increase of virological data on a global scale. Still, novel quantitative frameworks are needed to better visualize and analyze these data to extract meaningful information for response planning. Here we have shown that CoDA enables defining (sub)type composition trajectories, thus opening the door to state-of-the-art statistical tools for analyzing their spatiotemporal properties and forecasting their future evolution.

We have analyzed (sub)type composition at the global level, considering trajectories of annual compositions by country. This has revealed meaningful spatiotemporal patterns. We have characterized the evolution in time of the ensemble of trajectories by focusing on the extent of (sub)type mixing through the mixing score. We have highlighted years of strong dominance of a given (sub)type within each country – the strong dominance of A/H3H2 and A/H1N1pdm in almost all countries globally, respectively occurring in 2003 and 2009, and the spatial segregation of (sub)types occurring concomitantly to the COVID-19 pandemic. Previous studies on seasonal influenza disruption have primarily focused on changes in peak timing and wave severity (15, 40). Strong (sub)type dominance can have marked effects on the age profile of cases. For instance, mortality shifted toward young individuals during the 2009 A/H1N1pdm pandemic (42), likely due to residual immunity in older cohorts from early-life exposure to A/H1N1 viruses circulating before 1950 (42). In addition, a sudden reduction in viral diversity can have long-term consequences for influenza ecology, as exemplified by the disappearance of B\Yamagata since the COVID-19 pandemic (33, 43). The mixing score introduced here provides a concise metric for monitoring trends and comparing different locations, supporting surveillance and epidemiological interpretation. In this study, we focused on the impact of extreme events on (sub)type mixing. Beyond this, the score could be used as a response variable to test various predictors of dominance versus co-dominance, such as vaccine efficacy, vaccination coverage, population’s immune memory from past epidemics, and cross-immunity effects. This application would require compiling comprehensive, multi-country data on vaccine efficacy and coverage.

During a period free from disruptive events, we investigated the geographical drivers of (sub)type alternation. We found that pairs of countries with stronger air-travel connectivity, same epidemic periods, similar temperatures, and, to a lesser extent, relative humidity, exhibited more similar composition trajectories. While cross-immunity and waning immunity are known to be key drivers of (sub)type alternation (9–11, 13–16, 44), we found that geographical factors also played a significant role. Most notably, we identified an important role of air travel in homogenizing (sub)type composition across well-connected countries, especially in the north, leading to phase-locking of (sub)type alternation. International mobility was previously found to shape global phylogenies of the individual (sub)types (17, 18). Yet, no study addressed the role of this ingredient on global (sub)types co-existence so far. The emerging spatial structure was synthesized by the clustering analysis, which led to noteworthy results. Group I - i.e., Europe, North Africa, and West Asia - displayed a similar alternation of (sub)types, clearly distinguishable from the rest of the world. While such a strong similarity was noted before (19), here we show that it was due to the strong mobility coupling within the region compared to the rest of the world, together with the consistent climatic conditions and synchrony, confirming previous hypotheses (19). Furthermore, North America belonged to Group II and clustered with Eastern Asia in the finer clustering. Following H1N1pdm emergence, North America had a different alternation of A subtypes than Europe (15, 20). This was previously linked to the higher vaccination coverage in North America, which mitigated the circulation of A/H3N2, preventing it from dominating in the 2011/2012 season (15). While this factor may have played a role, we found that North America’s trajectories may have entered into phase with Eastern Asia’s ones due to the strong connectivity between the two regions. Finally, Australia and New Zealand had a different (sub)type alternation than northern hemisphere countries a few months later. Southern hemisphere countries are often regarded as sentinels for the approaching influenza season in northern countries (45–47). Still, the epidemiological evidence supporting this practice is contrasting (22, 48–50). For instance, the correlation between spatiotemporal indicators of Australia’s wave and the waves in the UK, the US, and Canada was limited (49, 50).

Grouping regions with similar influenza patterns is essential for surveillance. To this end, WHO defined 18 Influenza Transmission Zones (34). While previous studies evaluated the validity of the partition (19, 51, 52), only a few used (sub)type as a basis for their partition (19). Here, restricting the focus to (sub)type proportions, we found a subdivision broadly aligned with WHO ITZs, but coarser. Consistent with air travel being an important driving force of influenza composition, some clusters included noncontiguous yet strongly connected regions, e.g., North America and East Asia. This suggests the importance of considering mobility, together with climate and spatial proximity, when defining surveillance zones. Repeating this analysis after COVID-19 could reveal whether the patterns still hold and help to identify reliable priors for (sub)type circulation across regions.

The CoDa transformation enabled the use of state-of-the-art forecasting algorithms such as VAR and hierarchical VAR to predict the next year’s (sub)type composition. Trained on 2010–2019 data to avoid periods of disrupted circulation, VAR was limited by the short time series and performed no better than naive estimates. Still, hierarchical VAR improved performances by leveraging the geographical structure identified by the clustering. HVAR is directly applicable to time series expressed in log-ratio coordinates, whereas a similar analysis performed directly on percentage data would require defining and developing ad hoc algorithms. Interestingly, despite Group I countries being characterized by a marked A subtype alternation from one year to another, the *M2 H1-H3 alternation* model did not perform well, likely due to its inability to capture the trend of B proportion. Besides forecasting (sub)type proportions and the dominant (sub)type, we attempted to forecast binary outcomes, e.g., whether one specific (sub)type would be dominant (yes/no) or negligible (yes/no). Predictions were reasonably accurate for B negligibility (AUROC = 0.82), A/H3N2 negligibility (0.76), and dominance (0.70), especially for Group I countries. Predicting the dominance of A/H3N2 is important for preparedness since this is often associated with a high epidemic burden.

Epidemic forecasting is a central problem in infectious disease epidemiology. Consortia of modellers are dedicating a great effort to projecting seasonal influenza epidemics in the short and long term. In the context of these initiatives, a wide range of mechanistic and statistical models are being combined in ensemble modelling to forecast influenza incidence 1 to 4 weeks ahead or to identify possible evolution scenarios months ahead (53–57). This provides critical information for response planning. A different question regards the evolution of the influenza viruses. Several studies combine genetic and antigenic analyses to predict changes in the frequency of circulating clades for each (sub)type (58, 59). The identification of the clade more likely to be prevalent in the future informs the choice of vaccine composition, which needs to be done six months in advance. Here, we focused on a new task as we attempted to forecast (sub)type abundance by country one year ahead. We have framed the problem by introducing a range of models from the more naive to the more sophisticated. The (sub)type composition affects the expected epidemic size, the peak timing, and the age distribution of cases (6, 9, 11, 13). Therefore, its knowledge is precious for planning intervention measures. Predictions of next year’s (sub)type composition, if reliable, would provide a long horizon to plan the response, e.g., resource allocation, and targeted recommendation and vaccination. They could support scenario modelling and real-time incidence forecasts. For instance, probabilistic projections of (sub)type composition could be used to weigh possible scenarios. Knowing the dominant (sub)type at the start of the season would enable using statistical forecasting algorithms trained on the past seasons with the similar (sub)type frequency only, leading to higher prediction accuracy.

The HVAR model introduced here can be improved in the future. Restricting to the 50% best-classified countries improved the forecast for many observables. While identifying more predictable countries is informative for preparedness (60), this also suggests that incorporating information on other countries’ trajectories at different degrees according to the similarity could increase the quality of predictions. The HVAR model leveraged the clustering to coarsely account for the spatiotemporal pattern of subtype composition. More sophisticated models could explicitly account for the drivers of such a pattern. For instance, instead of relying on the geographical cluster, we could explicitly add the dependence on all other countries’ compositions, weighted by air travel distance. We could add to the VAR model terms for average temperature, humidity, and epidemic period. Antigenic drift and vaccination could be accounted for by incorporating their effect on dominance (44), leveraging recent works to forecast influenza evolution (61). Finally, new models could be designed to capture causal relationships between epidemics in different countries/years, e.g., through Bayesian networks (62).

CoDA is under-exploited in epidemiology. Analyses similar to the ones presented here could be applied to influenza (sub)type abundances at different spatiotemporal scales. For instance, weekly data would enable investigating within-season relative dynamics between A and B types (11, 63), addressing the shift between the two types’ waves, and investigating anomalous years (63). Within-country spatial coupling between compositions could also be investigated. More broadly, there is a variety of circulating viruses with distinct strains interfering with one another - SARS-CoV-2 variants, RSV types, HPV genotypes, and Dengue serotypes, to name a few. Percentage data are also used to monitor the co-circulation of bacterial strains. This is the case when studying the dynamics of Staphylococcus aureus strains with different antibiotic resistance profiles (63) or vaccine-targeted vs. non-targeted strains of Streptococcus pneumonia (64).

Our study is affected by limitations. Statistical forecasting models can be applied to periods of stable strain circulation, as was the case for 2010-2019. Following the COVID-19 pandemic, the global circulation of influenza viruses was highly perturbed, making any prediction more difficult. Reliable composition projections depend on FluNet data accurately representing countries’ viral compositions. Comparing sentinel and non-sentinel data showed a similarity between the two sources overall, although this varied across countries/years (Supplementary Information). Sentinel data are collected via a standardized protocol and are thus better interpretable. However, they contribute only to a small fraction of FluNet data at present (Supplementary Information). We have aggregated B/Victoria and B/Yamagata because lineage information for B samples was scarce - e.g., only 4 countries, among the ones satisfying the inclusion criteria, had at least 50% of B samples or at least 100 B samples, regardless of the percentage over the total, further subtyped in lineages for each year in the period 2010-2019. Still, the CoDA framework can be directly extended to the four (sub)type system whenever these data are available, thus enabling the investigation of the competitive dynamics between B lineages and their differential cross-immunity with A subtypes.

## Methods

### FluNet data

FluNet publishes weekly global data on influenza testing by country, reporting the number of samples tested and their results—positive or negative—categorized by influenza A subtypes, B lineages, and surveillance type (sentinel, non-sentinel, or undefined) (Supplementary Information) (1, 25). Throughout the Methods section, we will use for brevity H1 for A/H1N1 and A/H1N1pdm and H3 for A/H3N2. For each country, we determine influenza B infections by summing B\Yamagata, B\Victoria, and unspecified B samples. We grouped the pre-pandemic A/H1N1 and A/H1N1pdm. The un-subtyped influenza A samples were distributed between H1 and H3 according to the respective proportions of these subtypes among the subtyped A samples that week. If no A samples were subtyped that week, we looked at the proportions of H1 and H3 in the five weeks centred around the week or in the year. We aggregated influenza B, H1, and H3 samples over one year and calculated the respective percentages to define the relative abundance of the three influenza strains. Log-ratio transformations are not defined when any component equals zero. We thus replaced zeros with small values through a Geometric Bayesian-multiplicative treatment (Supplementary Information). The year considered in the analyses starts from week 17. As both the 2009 influenza and the COVID-19 pandemic began in spring, we set the year’s start in the spring to align southern temperate countries with northern temperate countries in terms of when the two pandemics affected the respective influenza seasons. The exact week was chosen by looking at global incidence profiles and minimising the risk of splitting an epidemic across years (Supplementary Information, Fig. S1). After verifying that (sub)types’ proportions were similar in sentinel and non-sentinel data (Supplementary Information), we aggregated the data regardless of the surveillance type. In all analyses, we discarded countries/years with fewer than 50 positive samples. In the sensitivity analysis, we tested alternative datasets using a different cut-off week for the definition of the year (37 instead of 17), and a threshold of 500 cases, instead of 50, for country inclusion.

### Definition of the mixing score

The *mixing score* is defined as the distance between the point in the log-ratio plane (u,v), representing the (sub)type composition, and the boundary of the co-dominance region, taken with a positive sign when the point is within that region and with a negative sign otherwise. The boundary of the co-dominance region in the simplex is identified by the points (B%, H1%, H3%) for which one component corresponds to exactly 50%. As an example, the points such that H1%=50%, are mapped by the *ilr* transformation into the coordinates (u, v(u)) such that

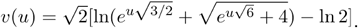

### Geographical predictors of annual trajectories of (sub)type compositions

We considered the 86 countries satisfying our inclusion criteria for the period 2010-2019. We defined the distances between pairs of country trajectories as the average Euclidean distance of the corresponding points in the (u,v) plane. We gathered the number of air-travel passengers between countries, countries’ average temperatures, relative humidities, and influenza seasons, among the northern hemisphere (NH), the southern hemisphere (SH), and year-round (YR). Then, we computed three continuous covariate matrices - the differences in temperatures and relative humidity, and the air travel distance between country pairs (65)-, and one categorical variable matrix describing epidemic synchrony, where country pairs were synchronous, i.e., NH-NH, SH-SH, or YR-YR, asynchronous, i.e,. NH-SH, or semi-synchronous, i.e., SH-YR or NR-YR. We then tested the association between trajectory distance and covariates using both univariable and multivariable analysis. In the Supplementary Information, we detail the data sources for covariates, the definition of covariates, the regression analyses, and the sensitivity analyses performed.

### Clustering of trajectories

We applied Ward’s linkage hierarchical clustering (66) to cluster countries with similar trajectories. Hierarchical clustering provides a hierarchy of nested partitions. We used the Silhouette coefficients (67) to choose the optimal number of clusters, i.e., the level of hierarchy. The optimal partition grouped countries into two groups. Robustness checks and sensitivity analyses (including alternative clustering algorithms) are described in the Supplementary Information.

### Forecasting of trajectories

We tested five forecasting algorithms to predict next year’s (sub)type composition in each country. We attempted to predict observables of different nature: (i) the multi-label categorical variable corresponding to the dominance state (dominance of B, H1, H3, or co-dominance); (ii) the (sub)type composition; (iii) the binary variables corresponding to the dominance (yes or no) and the negligibility (yes or no) of each (sub)type. Predictions of the three observables were associated with a quantification of the uncertainty in the prediction and a metric for prediction evaluation. However, not all observables, quantifications of uncertainty, and prediction evaluations were computable with all prediction methods. The five forecasting algorithms and the evaluation metrics used in the main paper were introduced in the Results section. We provide additional details hereafter. A complete summary of what can or cannot be computed across predicted observables and prediction methods is reported in the Supplementary Information, Table S1.

#### Algorithms for trajectory forecasting

We studied annual bivariate trajectories of compositions of the form (y_1_, …, y_T_), such that y_t_=(u,v)’_t_=*ilr*((B%, H1%, H3%)’_t_), with t=1,…,T and the single quote mark indicating transposed. We forecasted y_T+1_, i.e. the composition of 2017, 2018, or 2019, each time using the preceding period as the training set, i.e. (y_1_, …, y_T_) with y_1_ the composition in 2010. Below, we present the five forecasting methods.

*M1 past frequencies:* the dominance state was predicted to be the most frequently observed state during the preceding years if that state was unique. Otherwise, it was randomly chosen with uniform probability among the dominant states observed most often.

*M2 H1-H3 alternation:* the prediction for the y_T+1_ composition was ŷ_T+1_=(u,-v)’_T_, i.e., in percentage form, (B%_T+1_=B%_T_, H1%_T+1_=H3%_T_, H3%_T+1_=H1%_T_)’.

*M3 average composition:* the predicted composition was 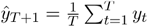, and the prediction’s confidence interval was given by the covariance matrix

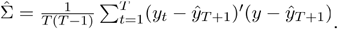

*M4 VAR:* assuming that the trajectory has been generated by a VAR process of lag *p*, then composition *t* can be written as a linear function of the previous *p* compositions: 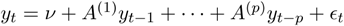, where *v* is the vector intercept, *A*^*(i*^) are 2×2 coefficient matrices, and is the Gaussian noise. This can be rewritten in matrix form as *Y = BZ + U* (36), where

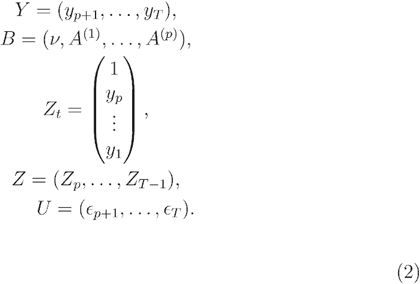

The resulting least squares estimator is 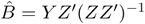. It follows that the prediction of the composition T+1 is 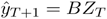, and the prediction’s confidence interval is estimated via the empirical covariance matrix corrected for short time series:

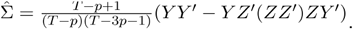

We only considered VAR processes with p=1, since for the shortest trajectories (prediction of 2017, with training set between 2010 and 2016), when p>1, the estimator of the covariance matrix is not defined.

*M5 HVAR:* the trajectories of the training set were clustered as described above in the Method section. For all three forecasted years, this yielded two groups that were almost identical to Group I and Group II obtained for the whole period 2010-2019 and discussed in the Results. For each cluster, we assumed that each trajectory followed a VAR process, such that the process for country *c* is: 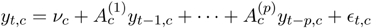. Similarly to the previous model, the coefficients can be encoded in the matrix 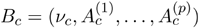, and the Gaussian noise in the matrix *U*_*C*_. Then, the hierarchical structure is imposed by assuming that the VAR processes for the trajectories in the group are similar. Specifically, we define *B*_*C*_ *= W + V*_*C*_, with *W* being the matrix of coefficients encoding the average behavior of the group, which is the same for all the trajectories in the group, and *V*_*C*_ being the coefficient matrix for the single trajectory adjustment. Moreover, we assume that elements in *W, V*_*C*_, *U*_*C*_ are independent random variables, sampled from distributions parametrized by some latent variables. All model equations are shown in the Supplementary Information. The model coefficients were optimized by maximum likelihood estimation using Monte Carlo methods. In particular, since for each coefficient it was possible to write the model’s likelihood conditional on the other coefficients, efficient estimation was performed using a Gibbs sampler. For the conditional likelihood distributions and the code to implement the Gibbs sampler, we followed (37), modifying their model to introduce the intercept terms. From the Monte Carlo chains, for each country, we sampled the distribution of predictions ŷ_T+1_. Then, from those samples, we computed the mean prediction and the covariance matrix for the confidence intervals. We tested HVAR models with p=1 and p=2.

#### Metrics for evaluating the goodness of predictions

To enable model comparison, the evaluation scores of different countries/years were averaged to obtain a single value for the model performance. We associated uncertainty with the forecast evaluation by computing a confidence interval with bootstrap. This corresponded to the Standard Error of the Mean of the evaluation metrics over 200 bootstrap samples of the set of predictions. The values reported in Table 1 correspond to the averages of the *Dominance State Accuracy* and *Energy Score* values for different countries/years, while the values in Table 2 are averages of the country/year *Area Under the Receiver Operating Characteristic Curve (AUROC)* scores.

To compare dominance state predictions (Table 1), we defined the *Dominance State Accuracy\* as the percentage of dominance states correctly predicted. To compare predictions of compositions (Table 1) we instead relied on probabilistic forecasting evaluation scores. Specifically, the *Energy Score* compares the composition *y* ∈ ℝ^2^ corresponding to the observation with the forecast distribution *F* defined by N samples *X*_*1*_,…, *X*_*N*_, *X*_*i*_ ∈ ℝ^2^. The formula for the *Energy Score* is

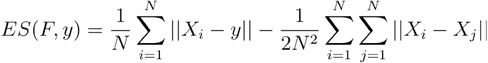

It is worth noting that this is a multivariate generalization of the more common *Continuous Rank Probability Score*, often used in epidemiology (38). Moreover, in the case of point forecast, it coincides with the *Mean Absolute Error*. Thus, it can also be used for evaluating the *M2 H1-H3 alternation* method, for which we do not have a confidence interval. The score is negatively oriented, so that it decreases when the forecast improves, and has a lower bound of zero for an ideal model. Furthermore, it is a *strictly proper* score, i.e., designed to be optimal only when the forecast distribution coincides with the true distribution of the observations (39). Binary predictions - i.e., dominance (yes/no) and negligibility (yes/no) of each (sub)type - were evaluated through the *Area Under the Receiver Operating Characteristic Curve (AUROC)* score (Table 2). It quantifies the overlap between the positive and negative classes and ranges from 0 to 1: 1 for an ideal model where the classes are perfectly separated, 0.5 for a random guess, and 0 for the worst possible model that misclassifies all observations.

For the *M5 HVAR* model, we tested lag=1 and lag=2. The results for the *M5 HVAR* model in all tables are obtained using the lag with the lowest *Energy Score* for each group of countries, i.e., lag=1 for Group II countries and lag=2 for Group I countries.

Other evaluation metrics were tested in the sensitivity analysis. They are described in the Supplementary Information, Table S1, while the results of these metrics are reported in the Supplementary Information, Tables S5 and S6.

## Data Availability

Code and data for reproducible analyses are available at
https://github.com/FrancescoBonacina/coupled-dynamics-flu-subtypes/ .

## Code and data availability

The global flight network data are commercially available from the International Air Transport Association (IATA, https://www.iata.org/en/contact-support). Analysis was implemented in R (version 4.3.2) and Python (version 3.8.5). Other than standard packages for data treatment, plots, and calculations (mainly the Python packages *pandas, matplotlib*, and *numpy*), we relied on the following packages for specific tasks:

- *zComposition 1*.*4*.*0-1* (R) for zero imputation in the pre-processing of compositions (68);
- *robCompositions 2*.*3*.*1* (R) for mapping compositions from the Simplex to the Euclidean space and back (69);
- *ternary* (python) for drawing ternary plots (70);
- *scipy 1*.*6*.*2* (python) for clustering analysis;
- *sklearn 1*.*3*.*2* (python) for clustering analysis and computation of some of the prediction evaluation scores (71);
- R code developed by Lu et al (37), on the basis of which we performed the VAR and HVAR predictions;
- *scoringRules 1*.*0*.*2* (R) for calculation of proper scores for probabilistic forecast evaluation (72).

Code and data for reproducible analyses are available at https://github.com/FrancescoBonacina/coupled-dynamics-flu-subtypes/.

## Acknowledgment

We thank Eugenio Valdano for the inputs on the analysis. This study was partially funded by EU grant 874850 MOOD to V.C. and is catalogued as MOOD 136. The contents of this publication are the sole responsibility of the authors and don’t necessarily reflect the views of the European Commission. Additional supporting funding was provided by the ANR project DATAREDUX(ANR-19-CE46-0008-03) to V.C.; the Municipality of Paris through the programme Emergence(s) to C.P. and F.B.; the BEHAVE-MOD project funded through the Cascade Open Call 2023-01 for Spoke 4 of the PNRR INF-ACT Grant to F.B.; Cariparo Foundation through the program Starting Package to C.P.; Department of Molecular Medicine through the program SID from BIRD funding to C.P.

## Author contribution

F.B., P.Y.B, V.C., O.L., M.T, and C.P. designed research; F.B. performed research; F.B, P.Y.B., and C.P. analyzed data; F.B., P.Y.B, V.C., O.L., M.T, and C.P. wrote the paper.

## References

1. GISRS, FluNet database - National Influenza Centres (NICs) of the Global Influenza Surveillance and Response System (GISRS) and World Health Organisation (WHO). (2022). Available at: https://www.who.int/tools/flunet [Accessed 8 April 2022].

2. P. Palese, T. T. Wang, Why Do Influenza Virus Subtypes Die Out? A Hypothesis. mBio 2, e00150–11 (2011).

3. K. L. Laurie, et al., Interval Between Infections and Viral Hierarchy Are Determinants of Viral Interference Following Influenza Virus Infection in a Ferret Model. J. Infect. Dis. 212, 1701–1710 (2015).

4. S. Caini, et al., Distribution of influenza virus types by age using case-based global surveillance data from twenty-nine countries, 1999-2014. BMC Infect. Dis. 18, 269 (2018).

5. A. Gagnon, E. Acosta, M. S. Miller, Age-Specific Incidence of Influenza A Responds to Change in Virus Subtype Dominance. Clin. Infect. Dis. 71, e195–e198 (2020).

6. M. C. Vieira, et al., Lineage-specific protection and immune imprinting shape the age distributions of influenza B cases. Nat. Commun. 12, 4313 (2021).

7. M. An der Heiden, U. Buchholz, Estimation of influenza-attributable medically attended acute respiratory illness by influenza type/subtype and age, Germany, 2001/02-2014/15. Influenza Other Respir. Viruses 11, 110–121 (2017).

8. S. Puzelli, et al., Co-circulation of the two influenza B lineages during 13 consecutive influenza surveillance seasons in Italy, 2004-2017. BMC Infect. Dis. 19, 990 (2019).

9. B. S. Finkelman, et al., Global Patterns in Seasonal Activity of Influenza A/H3N2, A/H1N1, and B from 1997 to 2005: Viral Coexistence and Latitudinal Gradients. PLoS ONE 2, e1296 (2007).

10. A. Suzuki, K. Mizumoto, A. R. Akhmetzhanov, H. Nishiura, Interaction Among Influenza Viruses A/H1N1, A/H3N2, and B in Japan. Int. J. Environ. Res. Public. Health 16, 4179 (2019).

11. E. Goldstein, S. Cobey, S. Takahashi, J. C. Miller, M. Lipsitch, Predicting the Epidemic Sizes of Influenza A/H1N1, A/H3N2, and B: A Statistical Method. PLOS Med. 8, e1001051 (2011).

12. K. L. Laurie, et al., Evidence for Viral Interference and Cross-reactive Protective Immunity Between Influenza B Virus Lineages. J. Infect. Dis. 217, 548–559 (2018).

13. W. Yang, E. H. Y. Lau, B. J. Cowling, Dynamic interactions of influenza viruses in Hong Kong during 1998-2018. PLOS Comput. Biol. 16, e1007989 (2020).

14. J. Truscott, et al., Essential epidemiological mechanisms underpinning the transmission dynamics of seasonal influenza. J. R. Soc. Interface 9, 304–312 (2011).

15. D. He, et al., Global Spatio-temporal Patterns of Influenza in the Post-pandemic Era. Sci. Rep. 5, 11013 (2015).

16. L. Gatti, et al., Cross-reactive immunity potentially drives global oscillation and opposed alternation patterns of seasonal influenza A viruses. Sci. Rep. 12, 8883 (2022).

17. T. Bedford, et al., Global circulation patterns of seasonal influenza viruses vary with antigenic drift. Nature 523, 217–220 (2015).

18. F. Parino, et al., Integrating dynamical modeling and phylogeographic inference to characterize global influenza circulation. PNAS Nexus 4, pgae561 (2025).

19. S. Caini, W. J. Alonso, C. E.-G. Séblain, F. Schellevis, J. Paget, The spatiotemporal characteristics of influenza A and B in the WHO European Region: can one define influenza transmission zones in Europe? Eurosurveillance 22, 30606 (2017).

20. P. Mook, et al., Alternating patterns of seasonal influenza activity in the WHO European Region following the 2009 pandemic, 2010-2018. Influenza Other Respir. Viruses 14, 150–161 (2020).

21. L. Zheng, et al., Global variability of influenza activity and virus subtype circulation from 2011 to 2023. BMJ Open Respir. Res. 10, e001638 (2023).

22. S. B. Choi, J. Kim, I. Ahn, Forecasting type-specific seasonal influenza after 26 weeks in the United States using influenza activities in other countries. PLOS ONE 14, e0220423 (2019).

23. J. Aitchison, The Statistical Analysis of Compositional Data. J. R. Stat. Soc. Ser. B Methodol. 44, 139–160 (1982).

24. P. Filzmoser, K. Hron, M. Templ, Applied Compositional Data Analysis: With Worked Examples in R (Springer International Publishing, 2018).

25. A. Flahault, et al., FluNet as a Tool for Global Monitoring of Influenza on the Web. JAMA 280, 1330–1332 (1998).

26. F. Chayes, On correlation between variables of constant sum. J. Geophys. Res. 1896-1977 65, 4185–4193 (1960).

27. J. J. Egozcue, V. Pawlowsky-Glahn, G. Mateu-Figueras, C. Barceló-Vidal, Isometric Logratio Transformations for Compositional Data Analysis. Math. Geol. 35, 279–300 (2003).

28. E. Ghedin, et al., Large-scale sequencing of human influenza reveals the dynamic nature of viral genome evolution. Nature 437, 1162–1166 (2005).

29. CDC, “Update: Influenza Activity --- United States and Worldwide, 2003--04 Season, and Composition of the 2004--05 Influenza Vaccine” (2004).

30. C. Fraser, et al., Pandemic Potential of a Strain of Influenza A (H1N1): Early Findings. Science 324, 1557–1561 (2009).

31. V. Dhanasekaran, et al., Human seasonal influenza under COVID-19 and the potential consequences of influenza lineage elimination. Nat. Commun. 13, 1721 (2022).

32. F. Bonacina, et al., Global patterns and drivers of influenza decline during the COVID-19 pandemic. Int. J. Infect. Dis. 128, 132–139 (2023).

33. Z. Chen, et al., COVID-19 pandemic interventions reshaped the global dispersal of seasonal influenza viruses. Science 386, eadq3003 (2024).

34. Influenza Transmission Zones. Available at: https://www.who.int/publications/m/item/influenza_transmission_zones [Accessed 18 April 2024].

35. Natural Earth, Natural Earth - Free vector and raster map data. Available at: https://www.naturalearthdata.com/ [Accessed 8 February 2024].

36. H. Lütkepohl, New introduction to multiple time series analysis: with … 36 tables, 1. ed., corr. 2. print (Springer, 2007).

37. F. Lu, Y. Zheng, H. Cleveland, C. Burton, D. Madigan, Bayesian hierarchical vector autoregressive models for patient-level predictive modeling. PLOS ONE 13, e0208082 (2018).

38. J. Bracher, E. L. Ray, T. Gneiting, N. G. Reich, Evaluating epidemic forecasts in an interval format. PLOS Comput. Biol. 17, e1008618 (2021).

39. T. Gneiting, A. E. Raftery, Strictly Proper Scoring Rules, Prediction, and Estimation. J. Am. Stat. Assoc. 102, 359–378 (2007).

40. M. A. Miller, C. Viboud, M. Balinska, L. Simonsen, The Signature Features of Influenza Pandemics — Implications for Policy. N. Engl. J. Med. 360, 2595–2598 (2009).

41. M. Lemaitre, F. Carrat, Comparative age distribution of influenza morbidity and mortality during seasonal influenza epidemics and the 2009 H1N1 pandemic. BMC Infect. Dis. 10, 162 (2010).

42. I. Skountzou, et al., Immunity to pre-1950 H1N1 influenza viruses confers cross-protection against the pandemic swine-origin 2009 A (H1N1) influenza virus. J. Immunol. Baltim. Md 1950 185, 1642–1649 (2010).

43. S. Caini, et al., Probable extinction of influenza B/Yamagata and its public health implications: a systematic literature review and assessment of global surveillance databases. Lancet Microbe 0 (2024).

44. A. C. Perofsky, et al., Antigenic drift and subtype interference shape A(H3N2) epidemic dynamics in the United States. eLife 13, RP91849 (2024).

45. D. F. Hughes, E. O’Neill, Flu season started early in Australia – countries in the northern hemisphere took note. The Conversation (2023). Available at: http://theconversation.com/flu-season-started-early-in-australia-countries-in-the-northern-hemisphere-took-note-207967 [Accessed 17 April 2024].

46. M. G. Baker, H. Kelly, N. Wilson, Pandemic H1N1 influenza lessons from the southern hemisphere. Eurosurveillance 14, 19370 (2009).

47. E. Harris, Southern Hemisphere Outcomes Support Flu Vaccine Effectiveness. JAMA 330, 1318–1319 (2023).

48. Y. Zhang, L. Yakob, M. B. Bonsall, W. Hu, Predicting seasonal influenza epidemics using cross-hemisphere influenza surveillance data and local internet query data. Sci. Rep. 9, 3262 (2019).

49. C. Viboud, et al., Influenza Epidemics in the United States, France, and Australia, 1972–19971. Emerg. Infect. Dis. 10, 32–39 (2004).

50. P. H. A. of Canada, Does the Australian influenza season predict the Canadian influenza season? CCDR 49(11/12). (2023). Available at: https://www.canada.ca/en/public-health/services/reports-publications/canada-communicable-disease-report-ccdr/monthly-issue/2023-49/issue-11-12-november-december-2023/does-australian-influenza-season-predict-canadian-influenza-season-2014-2020.html [Accessed 17 April 2024].

51. C. Chen, et al., The global region-specific epidemiologic characteristics of influenza: World Health Organization FluNet data from 1996 to 2021. Int. J. Infect. Dis. 129, 118–124 (2023).

52. W. J. Alonso, et al., A global map of hemispheric influenza vaccine recommendations based on local patterns of viral circulation. Sci. Rep. 5, 17214 (2015).

53. S. M. Mathis, et al., Evaluation of FluSight influenza forecasting in the 2021–22 and 2022–23 seasons with a new target laboratory-confirmed influenza hospitalizations. Nat. Commun. 15, 6289 (2024).

54. RespiCast | European Respiratory Diseases Forecasting Hub. Available at: https://respicast.ecdc.europa.eu/ [Accessed 18 April 2024].

55. CDC, About Flu Forecasting | CDC. (2023). Available at: https://www.cdc.gov/flu/weekly/flusight/how-flu-forecasting.htm [Accessed 6 January 2024].

56. Influcast. Influcast. Available at: https://influcast.org/ [Accessed 18 April 2024].

57. Home - Scenario model hub. Available at: https://scenariomodelinghub.org/ [Accessed 27 June 2024].

58. M. Luksza, M. Lässig, A predictive fitness model for influenza. Nature 507, 57–61 (2014).

59. R. A. Neher, T. Bedford, R. S. Daniels, C. A. Russell, B. I. Shraiman, Prediction, dynamics, and visualization of antigenic phenotypes of seasonal influenza viruses. Proc. Natl. Acad. Sci. U. S. A. 113, E1701–1709 (2016).

60. C. Viboud, A. Vespignani, The future of influenza forecasts. Proc. Natl. Acad. Sci. 116, 2802–2804 (2019).

61. J. Huddleston, T. Bedford, J. Chang, J. Lee, R. A. Neher, “Seasonal influenza circulation patterns and projections for February 2024 to February 2025” (Zenodo, 2024).

62. D. Heckerman, Bayesian Networks for Data Mining. Data Min. Knowl. Discov. 1, 79–119 (1997).

63. R. K. Borchering, et al., Anomalous influenza seasonality in the United States and the emergence of novel influenza B viruses. Proc. Natl. Acad. Sci. 118, e2012327118 (2021).

64. F. Blanquart, S. Lehtinen, C. Fraser, An evolutionary model to predict the frequency of antibiotic resistance under seasonal antibiotic use, and an application to Streptococcus pneumoniae. Proc. R. Soc. B Biol. Sci. 284, 20170679 (2017).

65. B. Faucher, et al., Drivers and impact of the early silent invasion of SARS-CoV-2 Alpha. Nat. Commun. 15, 2152 (2024).

66. J. H. Ward Jr., Hierarchical Grouping to Optimize an Objective Function. J. Am. Stat. Assoc. 58, 236–244 (1963).

67. P. J. Rousseeuw, Silhouettes: A graphical aid to the interpretation and validation of cluster analysis. J. Comput. Appl. Math. 20, 53–65 (1987).

68. J. Palarea-Albaladejo, J. A. Martín-Fernández, zCompositions — R package for multivariate imputation of left-censored data under a compositional approach. Chemom. Intell. Lab. Syst. 143, 85–96 (2015).

69. M. Templ, K. Hron, P. Filzmoser, “robCompositions: An R-package for Robust Statistical Analysis of Compositional Data” in Compositional Data Analysis, (John Wiley & Sons, Ltd, 2011), pp. 341–355.

70. M. Harper, et al., python-ternary: Ternary Plots in Python. (2015). 10.5281/zenodo.34938. Deposited 7 December 2015.

71. F. Pedregosa, et al., Scikit-learn: Machine Learning in Python. J. Mach. Learn. Res. 12, 2825–2830 (2011).

72. A. Jordan, F. Krüger, S. Lerch, Evaluating Probabilistic Forecasts with scoringRules. J. Stat. Softw. 90, 1–37 (2019).

