## Supplementary Information for "Characterization and forecast of global influenza (sub)type dynamics"

**Table of Contents**


**Additional methods** **3**
- Initial data processing 3
  - Definition of the influenza year 3
  - Treatment of zeros 4
  - Comparison between sentinel and non-sentinel data 4
- Geographical patterns: additional methods 5
  - Predictors tested for (sub)type composition trajectories 5
  - Univariable and multivariable analyses 6
- Forecast: additional methods 7
  - The Bayesian Hierarchical Vector AutoRegressive model (HVAR) 7
  - Summary of all quantities involved in the forecasting of trajectories 9

**Additional results** **14**
- Trends in (sub)type mixing: additional results 14
  - Geographical segregation of flu (sub)types in 2020-2021 14
- Geographical patterns: additional results 14
  - Geographical drivers of (sub)type composition trajectories 14
  - Grouping of countries up to the six-group partition 15
  - Differences between groups in terms of trajectories 16
  - Analysis of groups' characteristics 17
- Forecast: additional results 18
  - Insights into the estimated HVAR models 18

**Robustness checks and sensitivity analyses** **19**
- Definition of the different scenarios tested in the sensitivity analyses 19
- Alternative computations of the mixing score over time 20
- Geographical drivers of country similarities in (sub)type compositions under alternative scenarios 21
- Alternative trajectory clusterings 22
  - Alternative scenarios 22
  - Alternative clustering methods 22
- Alternative forecasting analyses and forecasting performance computations 23

**Additional Bibliography** **26**

### Additional methods

#### Initial data processing

Definition of the influenza year

We computed the percentage of weekly positive influenza viruses for each country and year with at least 50 classified cases between 2000 and 2024. Then, we averaged percentages over countries and years to obtain a single worldwide profile. This shows a large maximum in the winter (corresponding to the peak of northern hemisphere activity) and a small maximum in the summer (corresponding to the southern hemisphere peak), separated by local incidence minima in late April (week 17) and September (week 37). Fig. S1 shows that using either late April (rounded to May in the following and the main paper) or September as time breaks minimizes the chance of splitting seasonal epidemic waves occurring in the northern and southern hemispheres - note that splitting waves in the tropics is almost unavoidable as seasonal epidemics in the regions are more variable and irregular with heterogeneous behavior according to the country, e.g. year-round circulation, waves twice a year or every two years (1).

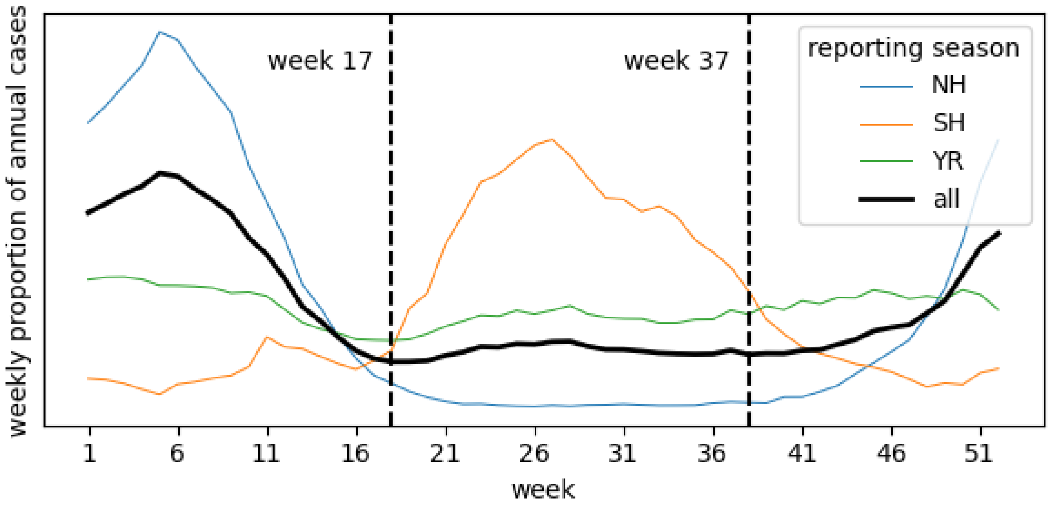


**Fig. S1 - Worldwide incidence.** Weekly proportion of annual cases averaged over time (years 2000 to 2024) and over all countries, as well as separately over countries with northern or southern hemisphere influenza seasons and those with year-round circulation.

The fact that both the 2009 A/H1N1 and COVID-19 pandemics started in spring led us to prefer week 17 as the start of the influenza year over week 37. In this way, for all countries, the first wave of the A/H1N1pdm pandemic was contained in 2009 and not split over two consecutive years. Similarly, the COVID-19 pandemic's effect on influenza circulation was more concentrated in 2020 and 2021 (2). A sensitivity analysis taking week 37 as the beginning of the year is reported in section “Robustness checks and sensitivity analyses” of the Supplementary Information.

Treatment of zeros

Neither the *isometric log-ratio transformatio*n defined in the section “Results” of the main paper nor the *additive log-ratio transformation*, tested in the sensitivity, is defined when any component equals zero. We thus first dealt with zeros, assuming that they were due to insufficient sample sizes, rather than a real absence of the virus. Specifically, we used a Geometric Bayesian-multiplicative treatment (3, 4) to replace the zero components with the Bayes estimator of a multinomial model, and then rescale all components to maintain their sum to one, keeping at the same time their ratios. The multinomial model assumes that the number of infections per (sub)type follows a multinomial distribution, whose parameters are distributed with a Dirichlet prior. This choice leads to a posterior distribution which is still a Dirichlet distribution, from which the Bayes estimator (i.e., the expectation of the posterior distribution) is easily computed.

Comparison between sentinel and non-sentinel data

Sentinel data are collected systematically by sentinel surveillance systems following a pre-defined protocol. On the other hand, non-sentinel data originate from heterogeneous sources, are collected in a non-standardized manner, and may include outbreak investigations, point-of-care testing, or other data sources (5). Classification in sentinels and non-sentinel was available for 314 countries/years, i.e., 14% of all countries/years satisfying our inclusion criteria.

The comparison between the log-ratio coordinates $u = \sqrt{\frac{2}{3}} \ln \frac{B\%}{\sqrt{H1\% * H3\%}}$ and $v = \sqrt{\frac{1}{2}} \ln \frac{H1\%}{H3\%}$ computed from sentinel and non-sentinel data is shown in Fig. S2. The line $y = x$ is shown as a guide to the eye. The two datasets are similar, with an intraclass correlation coefficient of 0.94, $p < 10^{-5}$(6).

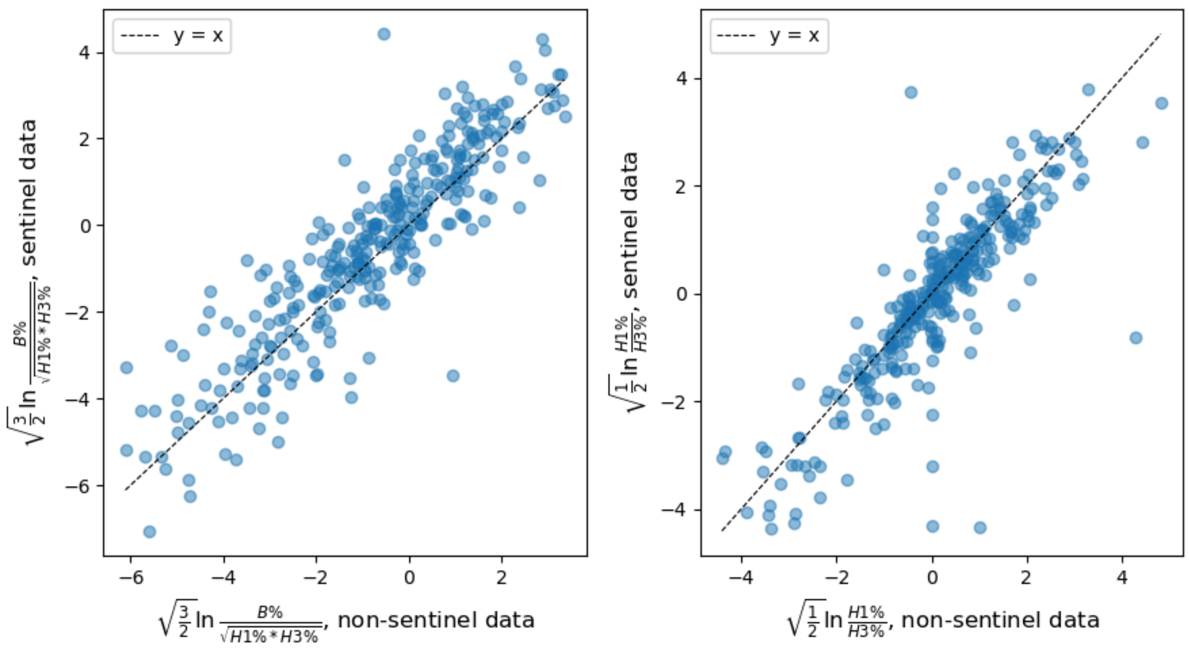


**Fig. S2 - Scatter plot of isometric log-ratio coordinates *u* (left) and *v* (right) computed from sentinel and non-sentinel data.** The *y=x* bisectors are shown as reference.

### Geographical patterns: additional methods

<u>Predictors tested for (sub)type composition trajectories</u>

To explain the similarity between countries in terms of (sub)type composition trajectories, we considered the intensity of flight connections between countries, whether they had synchronized epidemic seasons, and how similar the climatic conditions were. Specifically, the variables considered were defined as follows:

- *country-pair distances of (sub)type trajectories*: the response variable, defined as the average Euclidean distance between corresponding points of the trajectories in the (u, v) plane. It is a continuous variable.

- *country-pair synchrony of the epidemic seasons:* categorical covariate assuming values ‘synchronous’, ‘asynchronous, or ‘semi-synchronous’. The FluNet dataset provides information on the timing of influenza seasons in each country. Epidemic seasons typically peaking in alignment with the northern or southern hemisphere reporting periods are labeled as NH or SH, respectively, while countries with year-round circulation are marked as YR. Based on this classification, the synchronous pairs were the NH-NH, SH-SH, and YR-YR ones, the asynchronous were the NH-SH ones, and the semi-synchronous were NH-YR and SH-YR.

- *country-pair air-traffic distances:* continuous covariate. The strength of the flight connection between country pairs is defined based on the distance from country *i* to *j* introduced in (7, 8), i.e., the *ln(pax_ij / pop_i)*, where *pax_ij* represents the symmetrized volume of air passengers between *i* and *j* (data of 2019, from iata.org), and *pop_i* is the population size of country *i* (data from (9)). The expression was introduced to define an epidemic distance metric proportional to the average arrival time an epidemic in location *i* takes to be seeded in *j* (7, 10). Since this measure is not invariant under the permutation of *i* and *j*, we calculated a symmetric air-traffic distance by averaging the distances computed in both directions.
- *country-pair differences of temperature*: continuous covariate. We first computed country-wise average values of temperature from the ERA5-reanalysis dataset (11) and then country-pair differences. Country-wise temperatures were obtained by averaging monthly data over the period 2010-2019. For countries with epidemic seasons labeled as NH or SH, we considered only months that typically include the epidemic peak (>50% of the annual cases), namely January and February for the NH countries and June, July, and August for the SH countries. The ERA5 dataset provides climate data in a grid format; thus, to retrieve a unique value country-wise, we averaged the temperatures of the point of the grid in the proximity of the main cities within the country, weighted by the population size of the cities. This gave us the average temperature where most people live. As main cities, we considered all the cities with a population larger than 300K inhabitants (12), or the capital city, when all the cities in the country had a smaller population (13).
- *country-pair differences of relative humidity*: continuous covariate, defined analogously to the previous one.

As a preprocessing step, we applied monotonic transformations to the continuous variables to get more symmetrical distributions. Specifically, we log-transformed the (sub)type distances, we considered the square-root differences for temperature and relative humidity, and we left air-traffic distances unchanged. We then standardized all quantities.

<u>Univariable and multivariable analyses</u>

We tested the association between continuous covariates and the response variable one by one by using Mantel's test. Such a test is the equivalent of Spearman's correlation test for distance matrices, a kind of data for which the assumption of independence between observations does not hold (14). We investigated how the response variable differed among pairs of countries with varying epidemic synchrony by comparing distribution medians and applying permutation tests to determine statistical significance (10000 iterations).

As a second step, we included covariates in a multivariable linear mixed-effects model (see results reported in Table S2). Epidemic season synchrony was used as a fixed effect in the regression, with 'synchronous' observations as the reference group. To estimate the parameters' confidence intervals, we relied on a permutation procedure, as proposed in (15).

Note that for air travel, we had a single datum for the whole UK, whereas the FluNet data were disaggregated for England, Scotland, and Northern Ireland. Therefore, for this analysis, we considered values of the covariates computed over the whole UK, whereas for the (sub)type compositions, we attributed the value for England alone to the entire UK. This reduced the number of country/year observations from 86 to 84.

**Forecast: additional methods**

<u>The Bayesian Hierarchical Vector AutoRegressive model (HVAR)</u>

Let's consider a group of countries with similar trajectories. We assume that those country trajectories are generated by similar VAR processes, meaning that the coefficients of the VAR processes for the different countries are sampled from a common distribution, defined by some parameters to be estimated.

For each country *c*, the trajectory is of the form $(y_1, \ldots, y_T)_c$, with $y_t \in \mathbb{R}^k, t = 1, \ldots, T$. In our case $k = 2$, and the components of $y_t$ are the Euclidean coordinates $(u, v)'_t$. Thus, the VAR process of lag=*p* defined for country *c* is:

$$\begin{pmatrix} u \\ v \end{pmatrix}_{t,c} = \begin{pmatrix} \nu_u \\ \nu_v \end{pmatrix}_c + \begin{bmatrix} A_{uu}^{(1)} & A_{uv}^{(1)} \\ A_{vu}^{(1)} & A_{vv}^{(1)} \end{bmatrix}_c \begin{pmatrix} u \\ v \end{pmatrix}_{t-1,c} + \cdots + \begin{bmatrix} A_{uu}^{(p)} & A_{uv}^{(p)} \\ A_{vu}^{(p)} & A_{vv}^{(p)} \end{bmatrix}_c \begin{pmatrix} u \\ v \end{pmatrix}_{t-p,c} + \begin{pmatrix} \epsilon_u \\ \epsilon_v \end{pmatrix}_{t,c},$$

where $(\epsilon_u, \epsilon_v)'_{t,c}$ is Gaussian noise specified by the precision matrix $\Lambda \in \mathbb{R}^{2,2}$.

The same process can be written in a compact form. Following (16), let's define

$$
\begin{aligned}
Y_c &= (y_{p+1,c}, \dots, y_{T,c}), \\
B_c &= (\nu_c, A_c^{(1)}, \dots, A_c^{(p)}), \\
Z_{t,c} &= \begin{pmatrix} 1 \\ y_{p,c} \\ \vdots \\ y_{1,c} \end{pmatrix}, \\
Z_c &= (Z_{p,c}, \dots, Z_{T-1,c}), \\
U_c &= (\epsilon_{p+1,c}, \dots, \epsilon_{T,c}).
\end{aligned}
\tag{2}
$$

Then, the VAR process becomes $Y_c = B_c Z_c + U_c$.

At this point, the hierarchical structure is imposed by assuming that $B_c = W + V_c$, with $W$ being the matrix of coefficients encoding the average behavior of the group, that is the same for all the trajectories in the group, and $V_c$ being the coefficient matrix for the single trajectory adjustment. Assumptions on the distribution of the model coefficients $(W, V_c)$ and of the Gaussian noise $(U_c)$ are summarized hereafter. We adopt a notation very close to (17); refer there for additional details of the model.

- We assumed a multivariate normal prior for the country-level coefficients $V_c$, such that $vec(V_c)|\Theta \sim MVN(0, \Theta^{-1})$, with $diag(\Theta) = (\theta_1, \dots, \theta_{k(kp+1)})$. All $\theta_j$ coefficients, with $j$ ranging from 1 to $k(kp+1)$, have a unique Gamma prior $\Gamma(\alpha, \beta)$.
- For all countries, the Gaussian noise is defined by the same time-invariant precision matrix $\Lambda \in \mathbb{R}^{k,k}$. That is, $vec(U_c) \sim MVN(0, \mathbb{I}_T \otimes \Lambda^{-1})$, where $\mathbb{I}_T \otimes \Lambda^{-1}$ is a block diagonal matrix of size $(kT, kT)$, with blocks $\Lambda$ on the diagonal. Furthermore, $\Lambda$ is assumed to be a positive definite matrix, sampled from a Wishart distribution of parameters $S$, the $k$ x $k$ scale matrix, and $d$, the degrees of freedom.
- Elements of $W$ are sampled from a multivariate normal distribution centered at 0 and with precision matrix D: $vec(W)|D \sim MVN(0, D^{-1})$, where $vec(W)$ is a column vector containing the columns of $W$ stacked one after the other. The precision matrix $D$ is a diagonal matrix of size $k(kp+1)$, with entries determined by the parameters $\lambda_{2,j}$ and $\tau_j^2$ for $j = 1, \dots, k(kp+1)$, such that $diag(D) = (\lambda_{2,1} + \frac{1}{2\tau_1^2}, \dots, \lambda_{2,k(kp+1)} + \frac{1}{2\tau_{k(kp+1)}^2})$.

Coefficients $2\tau_j^2$ follow independent exponential distributions with rates $\frac{\lambda_{1,j}^2}{2\xi_j^2}$. The $\xi_j^2$ parameters are computed from the precision matrix $\Lambda^{-1}$ as in (17). The $\lambda_{1,j}$ and the $\lambda_{2,j}$ coefficients have $\Gamma(\mu_1, \nu_1)$ and $\Gamma(\mu_2, \nu_2)$ priors, respectively.

- In summary, we fitted the parameters $\theta_j, V_c, \Lambda, W, \lambda_{1,j}, \lambda_{2,j}, \tau_j^2$, with $j = 1, \ldots, k(kp+1)$. The hyper-parameters $(\mu_1, \nu_1, \mu_2, \nu_2, \alpha, \beta, S, d)$ are assumed to be known and, in practice, are sampled from uniform distributions at the initialization step of the Monte Carlo sampling.

For each parameter to be estimated, the likelihood of the model conditional on the other parameters can be written. As a result, the parameters can be estimated by Gibbs sampling. For conditional distributions and code for implementing the Gibbs sampler, we followed (17). The only difference is that their model does not contain the intercept term.

Summary of all quantities involved in the forecasting of trajectories

In Table S1, we summarize the computable quantities for each forecasted observable and forecasting method. We also detail the alternative metric used for forecast evaluation.

Of note, alternative metrics used for evaluating probabilistic predictions of (sub)type compositions were the *Dawid-Sebastiani Score* and the *Variogram Score*. Resuming the notation used in the Methods, we use $y \in \mathbb{R}^2$ to refer to the observed composition and *F* to refer to the forecast distribution. *F* is defined by N samples $X_1, \ldots, X_N$, $X_i \in \mathbb{R}^2$ of the posterior distribution, for the *M5 HAVR* method, or by the estimated mean $\mu_X$ and covariance matrix $\Sigma_X$ for the *M3 average* and *M4 VAR* methods.

The *Dawid-Sebastiani Score* is the multivariate generalization of the *Logarithmic Score* (18), which is commonly used in epidemiology (19). It is defined as a function of the mean $\mu_X$ and covariance matrix $\Sigma_X$ of the forecast distribution F:
$DS(F, y) = \log(|\Sigma_X|) + (y - \mu_X)'\Sigma_X^{-1}(y - \mu_X)$.

The *Variogram Score* is more suitable for evaluating the correct or incorrect estimation of the correlations between components of the multivariate quantity (20), and it is defined as

$VS^p(F, y) = \sum_{i=1}^{d} \sum_{j=1}^{d} w_{i,j}(|y^{(i)} - y^{(j)}|^p - \frac{1}{N}\sum_{k=1}^{N} |X_k^{(i)} - X_k^{(j)}|^p)^2$. We considered p=0.5 and p=1 with constant weights $w_{i,j} = 1$ as standard choices. These two scores are negatively-oriented - i.e., smaller values indicate better performances - and proper scores.

| Quantity | | | Is the quantity computable for the specific forecasting method? | | | | |
|---|---|---|---|---|---|---|---|
| | **Notation** | **Description** | ***M1*** | ***M2*** | ***M3*** | ***M4*** | ***M5*** |
| **Observable 1 - multi-label categorical variable corresponding to the dominance state:** $s_{OBS}$, the observed dominance state, among four possible states: B, H1, H3, and co-dominance. They represent the dominance of one (sub)type (≥50% of cases) or the co-dominance of the three (sub)types, respectively. | | | | | | | |
| Prediction | $\hat{s}$ | The predicted dominance state among B, H1, H3, and co-dominance. It is defined by looking at the dominance region where $(\widehat{u}, \widehat{v})$ falls. | ✓ | ✓ | ✓ | ✓ | ✓ |
| Prediction uncertainties | $\{\hat{s}_i\}$ | Set of dominance states computed by looking at the dominance regions in which the samples $\{(\widehat{u}, \widehat{v})_i\}$ fall. The proportion of samples $\{\hat{s}_i\}$ belonging to each dominance region (B, H1, H3, or co-dominance) gives an estimate of the probability of observing such a dominance state. For method *M1*, the samples $\{\hat{s}_i\}$ correspond to the dominance states of the past years. | ✓ | X | ✓ | ✓ | ✓ |
| Evaluation scores | Dominance state accuracy (main metric) | The proportion of correct predictions, when comparing the observed ($s_{OBS}$) with the predicted ($\hat{s}$) dominance states. | ✓ | ✓ | ✓ | ✓ | ✓ |
| | Average Precision (from PR-curve) (sensitivity analysis) | It is a score used to summarize a precision-recall curve for binary classification, computed by comparing the observed binary event with the estimated probability of obtaining the event. In the case of multilabel classification, several ways to trace the problem back to a binary problem exist. Here, we averaged scores from all four definitions implemented in the Python package *sklearn.metrics* (21). In our case, we compared the observed dominance states ($s_{OBS}$) with the probabilities of occurrence of each dominant state estimated by the percentage of samples $\{\hat{s}_i\}$ belonging to each state (B, H1, H3, or co-dominance). | ✓ | X | ✓ | ✓ | ✓ |
| | AUROC (sensitivity analysis) | The Area Under the Receiver Operating Characteristic (ROC) Curve. It is a score used to summarize a ROC curve for binary | ✓ | X | ✓ | ✓ | ✓ |

| Quantity | | | Is the quantity computable for the specific forecasting method? | | | | |
|---|---|---|---|---|---|---|---|
| | Notation | Description | *M1* | *M2* | *M3* | *M4* | *M5* |
| | | classification. It is computed by comparing the observed binary event with the estimated probability of obtaining the event. In the case of multilabel classification, several ways to trace the problem back to a binary problem exist. Here we averaged scores from four definitions implemented in the Python package *sklearn.metrics* (21); specifically, we considered the *One-vs-Rest* and *One-vs-One* binarization procedures, and the *macro* and the *weighted* type of averages. In our case, we compared the observed dominance states ($s_{OBS}$) with the probabilities of occurrence of each dominant state estimated by the percentage of samples $\{\hat{s}_i\}$ belonging to each state (B, H1, H3, or co-dominance). | | | | | |
| **Observable 2 - (sub)type composition:** $(B\%,H1\%,H3\%)_{OBS}$ ⇔ $(u,v)_{OBS}$, the observed (sub)type composition. | | | | | | | |
| Prediction | $(\hat{B}\%,\hat{H}1\%,\hat{H}3\%)$ ⇔ $(\widehat{u,v})$ | The predicted composition one year ahead. It corresponds to the center of the forecast distribution. | X | ✓ | ✓ | ✓ | ✓ |
| Prediction uncertainties | $\hat{\Sigma}_{(u,v)}$ ⇔ $\{(\widehat{u,v})_i\}$ | The prediction uncertainties are given by the estimated covariance matrix for methods *M3* and *M4*, and by a set of compositions sampled from the forecast distribution for method *M5*. The two quantities express the same information since from the empirical samples one can estimate the covariance matrix (and vice-versa). Then, the forecast distribution is approximated as the bivariate normal distribution $N((\widehat{u,v})', \hat{\Sigma}_{(u,v)})$. | X | X | ✓ | ✓ | ✓ |
| Evaluation scores | ES (main metric) | The Energy Score (18). It is computed by comparing the *true* composition $(u,v)_{OBS}$ with samples from the forecast distribution $\{(\widehat{u,v})_i\}$. It coincides with the Mean Absolute Error for point forecasts. | X | ✓ | ✓ | ✓ | ✓ |
| | DSS (sensitivity analysis) | The Dawid-Sebastiani Score (18). It is computed by comparing the *true* composition $(u,v)_{OBS}$ with the forecast distribution, defined by its empirical mean $(\widehat{u,v})$ and its empirical covariance matrix $\hat{\Sigma}_{(u,v)}$. The Dawid-Sebastiani Score is not defined when only point forecasts are available. | X | X | ✓ | ✓ | ✓ |
| | VS05 (sensitivity analysis) | The Variogram Score of order 0.5 (20). It is computed by comparing the *true* composition $(u,v)_{OBS}$ with samples from the forecast | X | X | ✓ | ✓ | ✓ |

| Quantity | | | Is the quantity computable for the specific forecasting method? | | | | |
|---|---|---|---|---|---|---|---|
| | Notation | Description | *M1* | *M2* | *M3* | *M4* | *M5* |
| | | distribution $\{(\widehat{u},\widehat{v})_i\}$. The Variogram Score is not defined for point forecasts. | | | | | |
| | VS1 (sensitivity analysis) | The Variogram Score of order 1 (20). It is computed by comparing the *true* composition $(u,v)_{OBS}$ with samples from the forecast distribution $\{(\widehat{u},\widehat{v})_i\}$. The Variogram Score is not defined for point forecasts. | X | X | ✓ | ✓ | ✓ |
| **Observable 3 - binary variables corresponding to the dominance (yes or no) of each (sub)type:** $B\%_{OBS} \geq 50\%$, binary variable. It is equal to 1 if the observed percentage of B infections is at least 50%, 0 otherwise. The same quantities are similarly defined for H1 and H3. For simplicity, the notation below is written for (sub)type B. | | | | | | | |
| Prediction | $\hat{B}\% \geq 50\%$ | Binary variable: 1 if the predicted percentage of B infections is at least 50%, 0 otherwise. | X | ✓ | ✓ | ✓ | ✓ |
| Prediction uncertainties | $\frac{1}{n}\sum_{i=1}^{n}(\hat{B}\%_i \geq 50\%)$ | The proportion of $\{(\widehat{u},\widehat{v})_i\}$ compositions $\frac{1}{n}\sum_{i=1}^{n}(\hat{B}\%_i \geq 50\%)$, sampled from the forecast distribution (with *n* the number of samples) for which the percentage of B infections is at least 50%. For method *M1*, such a proportion is the fraction of past years in which B was dominant. | ✓ | X | ✓ | ✓ | ✓ |
| Evaluation scores | AUROC (main metric) | The Area Under the Receiver Operating Characteristic (ROC) Curve. It is a score used to summarize a ROC curve for binary classification. It is computed by comparing the observed dominance of B ($B\%_{OBS} \geq 50\%$) with the probability of observing at least 50% of B infections ($\frac{1}{n}\sum_{i=1}^{m}(\hat{B}_i \geq 50\%)$). | ✓ | X | ✓ | ✓ | ✓ |
| | Average Precision (from PR-curve) (sensitivity analysis) | It is a score used to summarize a precision-recall curve for binary classification. It is computed by comparing the observed dominance of B ($B\%_{OBS} \geq 50\%$) with the estimated probability of observing at least 50% of B infections ($\frac{1}{n}\sum_{i=1}^{n}(\hat{B}\%_i \geq 50\%)$). | ✓ | X | ✓ | ✓ | ✓ |
| | Precision (sensitivity analysis) | The ratio TP/(TP+FP). It is used for evaluating classification tasks, with TP being the number of true positives and FP the number of false positives. TP and FP are computed by comparing the observed dominance of B ($B\%_{OBS} \geq 50\%$) with the predicted dominance ($\hat{B}\% \geq 50\%$). | X | ✓ | ✓ | ✓ | ✓ |

| Quantity | | | Is the quantity computable for the specific forecasting method? | | | | |
|---|---|---|---|---|---|---|---|
| | **Notation** | **Description** | ***M1*** | ***M2*** | ***M3*** | ***M4*** | ***M5*** |
| **Observable 4 - binary variables corresponding to the negligibility (yes or no) of each (sub)type:** B%$_{OBS}$ < 10%, binary variable. It is equal to 1 if the observed percentage of B infections is below 10%, 0 otherwise. The same quantities are similarly defined for H1 and H3. For simplicity, the notation below is written for (sub)type B. | | | | | | | |
| Prediction | $\hat{B}$% < 10% | Binary variable: 1 if the predicted percentage of B infections is below 10%, 0 otherwise. | X | ✓ | ✓ | ✓ | ✓ |
| Prediction uncertainties | $\frac{1}{n}\sum_{i=1}^{n}(\hat{B}\%_i < 10\%)$ | The proportion of $\{(\widehat{u}, \widehat{v})_i\}$ compositions $\frac{1}{n}\sum_{i=1}^{n}(\hat{B}\%_i < 10\%)$, sampled from the forecast distribution (with *n* the number of samples), for which the percentage of B infections is less than 10%. For method *M1*, such a proportion is the fraction of past years in which B was negligible. | ✓ | X | ✓ | ✓ | ✓ |
| Evaluation scores | AUROC (main metric) | The Area Under the Receiver Operating Characteristic (ROC) Curve. It is a score used to summarize a ROC curve for binary classification. It is computed by comparing the observed negligibility of B (B%$_{OBS}$ < 10%) with the probability of observing less than 10% of B infections ($\frac{1}{n}\sum_{i=1}^{m}(\hat{B}_i < 10\%)$). | ✓ | X | ✓ | ✓ | ✓ |
| | Average Precision (from PR-curve) (sensitivity analysis) | It is a score used to summarize a precision-recall curve for binary classification. It is computed by comparing the observed negligibility of B (B%$_{OBS}$ < 10%) with the estimated probability of observing less than 10% of B infections ($\frac{1}{n}\sum_{i=1}^{n}(\hat{B}\%_i < 10\%)$). | ✓ | X | ✓ | ✓ | ✓ |
| | Precision (sensitivity analysis) | The ratio TP/(TP+FP). It is used for evaluating classification tasks, with TP being the number of true positives and FP the number of false positives. TP and FP are computed by comparing the observed negligibility of B (B%$_{OBS}$ < 10%) with the predicted negligibility ( $\hat{B}$% < 10%). | X | ✓ | ✓ | ✓ | ✓ |

**Table S1 - Computable quantities for each forecasting method.** In the table, we used for brevity H1 for A/H1N1 and A/H1N1pdm and H3 for A/H3N2.

### Additional results

#### Trends in (sub)type mixing: additional results

<u>Geographical segregation of flu (sub)types in 2020-2021</u>

From May 2020 to April 2021, only 31 countries reported at least 50 cases of influenza. Spatial segregation of (sub)types was unusually strong during that period (Fig. S3). We recovered a geographical distribution of (sub)type consistent with (22). In 13 countries, one (sub)type was responsible for more than 75% of the cases. A/H3N2 was nearly the sole circulating influenza strain in seven countries of South-East Asia (India, Nepal, Bangladesh, Cambodia, Vietnam, Laos, and Timor-Leste). Influenza B accounted for more than 80% of flu cases in five countries (Saudi Arabia, Haiti, China, Ethiopia, and Afghanistan), and it was the dominant strain in some American countries and Pakistan. A/H1N1 circulated mainly in Togo, Niger, South Korea.

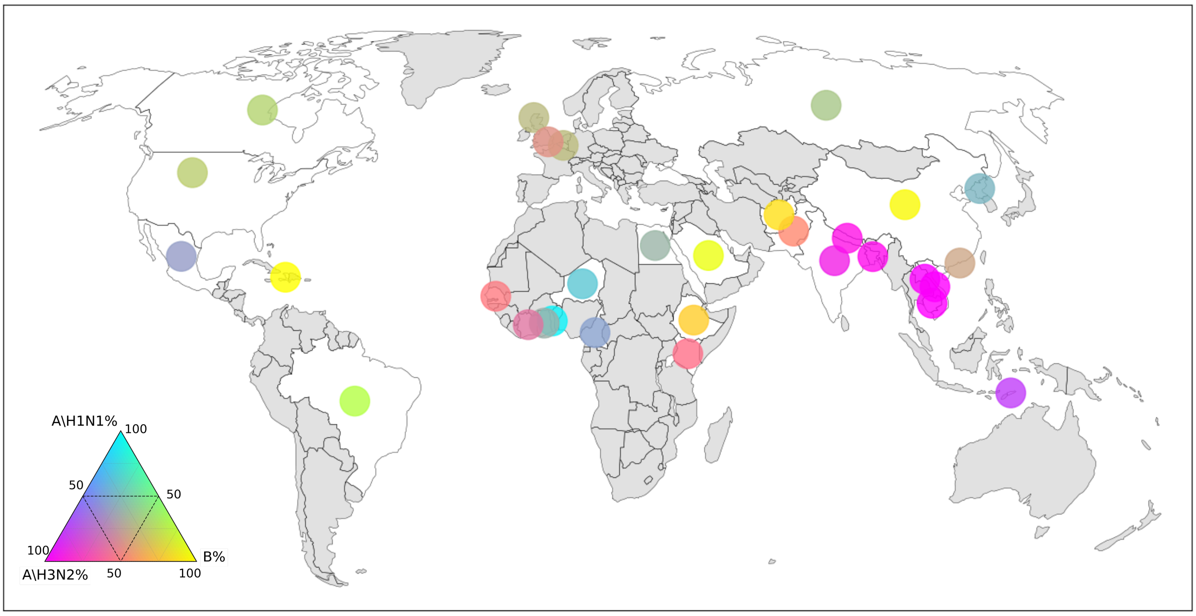


**Fig. S3 - Relative abundances of influenza (sub)types from May 2020 to April 2021.** Countries in gray did not report a minimum of 50 classified cases of influenza in the period. For the other countries, the circle's color represents the relative abundance of the (sub)types according to the scale defined by the ternary diagram at the bottom left.

#### Geographical patterns: additional results

<u>Geographical drivers of (sub)type composition trajectories</u>

In Table S2, we provide the results of the regression analysis to test the association between the distance between composition trajectories and synchrony of the epidemic seasons (‘synchronous’, ‘asynchronous, or ‘semi-synchronous’), air-traffic distance, difference of temperature, and difference of relative humidity.

| Variable | coefficient | p-value |
|---|---|---|
| Intercept | -0.127 | 0.0002 |
| Synchrony (semi-synchronous) | 0.143 | 0.012 |
| Synchrony (asynchronous) | 0.765 | 0 |
| Air-traffic distance | 0.298 | 0 |
| Difference in temperature | 0.241 | 0 |
| Difference in relative humidity | 0.101 | 0 |

**Table S2 - Coefficient of the multivariable regression for predicting the distance between countries’ (sub)type composition trajectories.**

### Grouping of countries up to the six-group partition

In Fig. S4, we report a diagram of the grouping of countries identified by Ward's hierarchical clustering algorithm, developed up to the six-groups partition. The six groups are also compared with the Influenza Transmission Zones (ITZ) defined by the WHO. ITZs tend to belong entirely to one group of the two-group partition. Similarly, when considering the six-group partition, they split into different subgroups only in a few cases.

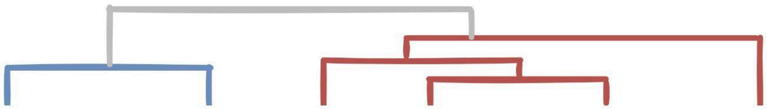

| Clustering of (sub)type composition trajectories | Group I (41 countries) | | Group II (45 countries) | | | |
|---|---|---|---|---|---|---|
| **WHO Influenza transmission Zones** | **Europe (36)** | **Western Asia (5)** | **Qatar (1)** | **Temperate North (7)** | **Tropics (31)** | **South and Central America (6)** |
| Northern Europe | DNK, EST, FIN, ISL, IRL, LVA, LTU, NOR, SWE, X09, X11, X12 | | | | | |
| South West Europe | ALB, AUT, BEL, HRV, FRA, DEU, GRC, ITA, LUX, NLD, PRT, SRB, SVN, ESP, CHE | | | | | |
| Eastern Europe | BLR, BGR, CZE, HUN, ROU, RUS, SVK | UKR | | | | |
| Northern Africa | EGY | TUN | | | | |
| Western Asia | ISR | JOR, TUR | QAT | | OMN | |
| Southern Asia | | IRN | | | BGD, IND, LKA | |
| South East Asia | | | | | KHM, IDN, LAO, PHL, SGP, THA, VNM | |
| Oceania Melanesia and Polynesia | | | | | AUS, NZL | |
| Central Asia | | | | | KAZ | |
| Middle Africa | | | | | CMR | |
| Eastern Africa | | | | | MDG, TZA | |
| Western Africa | | | | | CIV, GHA, SEN | |
| Southern Africa | | | | | ZAF | |
| Tropical South America | | | | | BOL, BRA, COL, ECU, GUF, PER | |
| Central America | | | | | CUB, GTM, HND | CRI, SLV, MEX, NIC |
| Temperate South America | | | | | PRY | ARG, CHL |
| Eastern Asia | | | | CHN, HKG, JPN, MNG, KOR | | |
| Northern America | | | | CAN, USA | | |

**Fig. S4 - Hierarchical clustering of country trajectories compared with the W.H.O. Influenza Transmission Zones.** The columns correspond to the country groups identified by Ward's linkage hierarchical clustering up to the six-groups partition. The nested structure of the clustering is specified by the dendrogram at the top. The rows designate the Influenza Transmission Zones of the WHO. The 86 countries considered in the analysis are identified by their three-letter ISO codes. X09, X11, and X12 correspond to England, Northern Ireland, and Scotland, respectively. We used blue and red to show the correspondence between our clustering and the WHO ITZs. The three countries in purple are the only ones for which the two groupings do not match.

### Differences between groups in terms of trajectories

We compare average trajectories for the different country groups identified by the hierarchical clustering to highlight the main differences in patterns of (sub)type alternation (Fig. S5). In particular, we compare Group I vs. Group II (excluding Qatar, since it has a specific stand-alone behavior) and the two subgroups of Group I (*Europe* and *Western Asia*) with the three individual subgroups that compose Group II - namely *South and Central America, Temperate North,* and *Tropics*. The results indicate that tropical countries (*Tropics*) experienced very limited alternation of (sub)types during 2010-2019, compared to the other groups. On the other hand, the alternations of (sub)types in *South and Central America* and *Temperate North* countries, although pronounced, were not synchronized with each other and with countries from *Europe* and *Western Asia*.

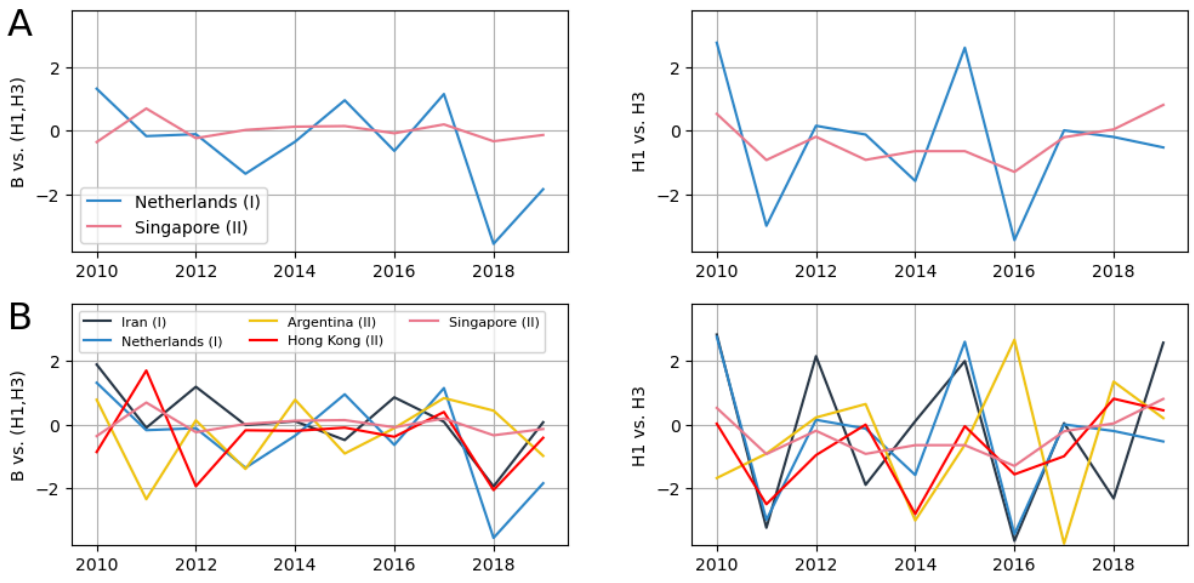


**Fig. S5 - Medoid trajectories of (sub)type alternation by country groups.** The bivariate trajectories of the medoid - i.e., the most central trajectories - of the different groups and subgroups are plotted with solid lines. A) Comparison of trajectories for the Netherlands and Singapore, respectively, medoids of Group I and Group II in the two-group partition. B) Comparison of the trajectories of Iran, the Netherlands, Argentina, Hong Kong, and Singapore, medoids of the five groups of the six-group partition. Qatar is excluded as it has a specific stand-alone behaviour that is not representative of other countries. In the figure, H3 is used to indicate H3N2, and H1 is used to indicate A/H1N1 before 2009, and A/H1N1pdm after 2009.

### Analysis of groups' characteristics

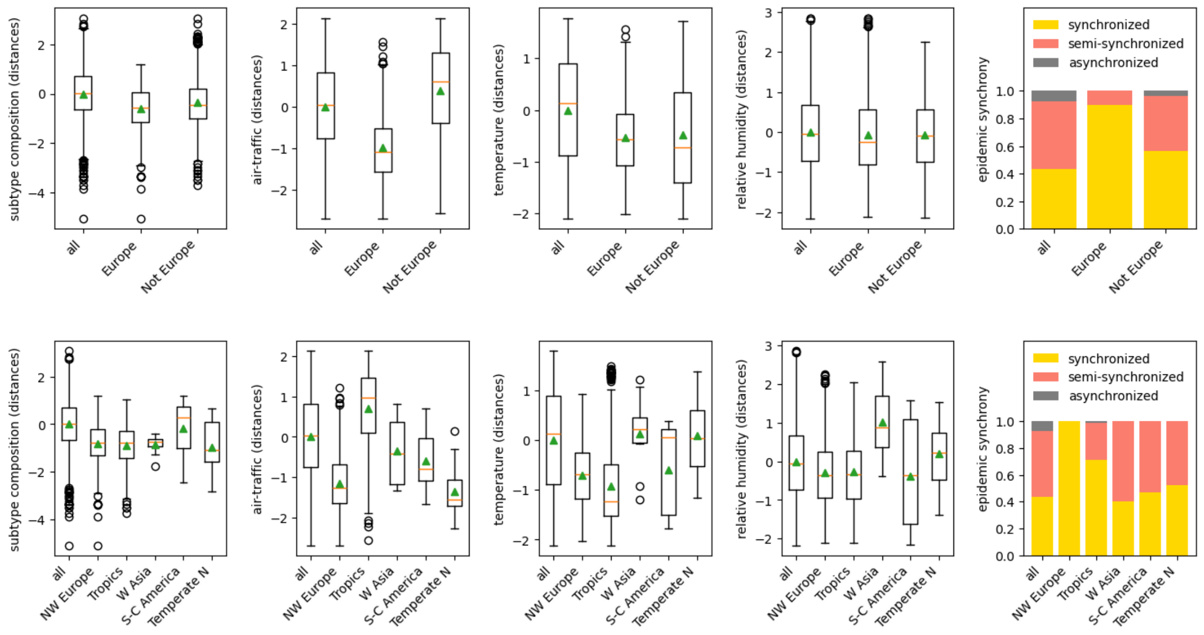


**Fig. S6 - Distribution of covariates for the groups of countries defined by the two-groups partition (top panel) and the six-groups partition (bottom panel).** For each variable, the differences between all countries and between countries in the same group are compared (x-axis). Before computing the differences, variables have been processed and standardised as explained in the section “Predictors tested for (sub)type composition trajectories” of the Supplementary Information.

### Forecast: additional results

#### Insights into the estimated HVAR models

In Fig. S7, we report the distributions of the VAR coefficients estimated through the *M5 HVAR* model for Group I and Group II trajectories (left and right panels, respectively). For each group, we consider the lag which minimizes the average Energy Score over all countries within the group and all predicted years (2017, 2018, 2019), namely lag=2 for Group I and lag=1 for Group II. The VAR process for each country *c* of Group I is defined as $\begin{pmatrix} u \\ v \end{pmatrix}_{t,c} \simeq \begin{pmatrix} \nu_u \\ \nu_v \end{pmatrix}_c + \begin{bmatrix} A^{(1)}_{uu} & A^{(1)}_{uv} \\ A^{(1)}_{vu} & A^{(1)}_{vv} \end{bmatrix}_c \begin{pmatrix} u \\ v \end{pmatrix}_{t-1,c} + \begin{bmatrix} A^{(2)}_{uu} & A^{(2)}_{uv} \\ A^{(2)}_{vu} & A^{(2)}_{vv} \end{bmatrix}_c \begin{pmatrix} u \\ v \end{pmatrix}_{t-2,c}$. The same process, without the last term, was used to model Group II trajectories.

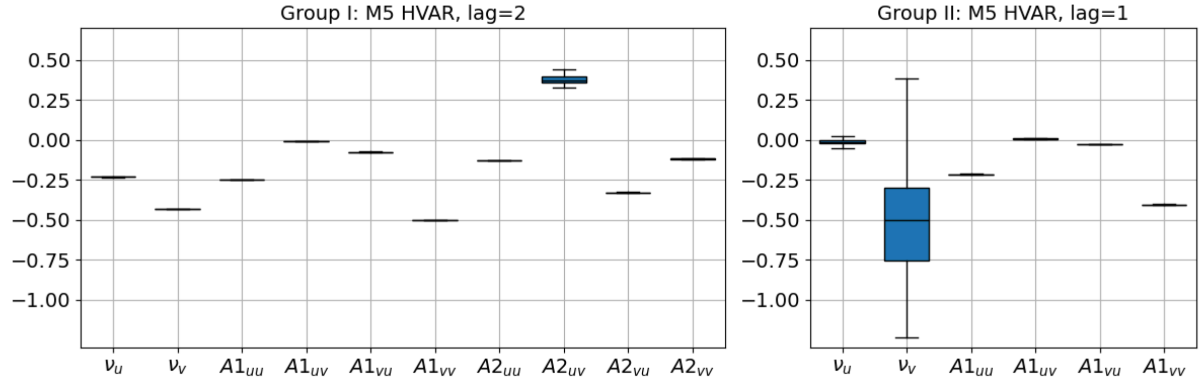


**Fig. S7 - Distributions of coefficients of the HVAR models estimated for Group I (lag=2) and Group II (lag=1) trajectories.**

The VAR processes estimated for each country are similar to each other. The offsets ($(\nu_u, \nu_v)'$) of the processes estimated for Group I countries signal that the co-dominance of (sub)types or the dominance of A/H3N2 are the most likely situations, with A/H3N2 abundance close to 50%. This is in line with the fact that overall A/H3N2 is the most circulating subtype (23, 24) due to its fast mutation rate and higher transmissibility (25, 26). The same tendency holds for Group II countries, but with a higher mixing of (sub)types on average (indicated by the larger value of $\nu_u$ ) and a higher country-by-country variability.

The other coefficients in the models rule the viruses' alternation and are robust across countries. They are almost all negative, indicating that the (sub)types tend to alternate from one year to the next. This is consistent with the cross-immunity effect described in the literature (27–31).

#### Robustness checks and sensitivity analyses

#### Definition of the different scenarios tested in the sensitivity analyses

The alternative scenarios tested in the sensitivity analyses are summarized in Table S3 and described in detail below.

| | **main analysis** | **sensitivity analyses** | | | |
|---|---|---|---|---|---|
| **scenario** | **base scenario** | **alr coordinates** | **fall-year** | **500+ annual cases** | **best classified countries** |
| **no. of selected countries** | 86 | 86 | 88 | 29 | 43 |
| | **criteria defining the scenarios:** | | | | |
| **minimum no. of annual cases** | 50 | 50 | 50 | 500 | 50 |
| **1st week of the influenza year** | 17 | 17 | 37 | 17 | 17 |
| **log-ratio transformation** | *ilr* | *alr* | *ilr* | *ilr* | *ilr* |
| **period *** | 2010 to 2019 | 2010 to 2019 | 2010 to 2018 | 2010 to 2019 | 2010 to 2019 |
| **additional filter to select trajectories with stable clustering** | FALSE | FALSE | FALSE | FALSE | TRUE |
| | **the scenario has been tested in the following analyses:** | | | | |
| **mixing score** | ✓ | ✓ | X | X | X |
| **clustering** | ✓ | ✓ | ✓ | ✓ | X |
| **prediction of country-pair distances of (sub)type compositions** | ✓ | ✓ | ✓ | ✓ | X |
| **forecast** | ✓ | ✓ | ✓ | ✓ | ✓ |

* year *y* refers to the influenza year between years *y* and *y+1*

**Table S3 - Summary of alternative scenarios considered for the sensitivity analyses.**

Alternative log-ratio transformation: In the main paper, we used the isometric log-ratio (ilr) transformation to map the composition vectors belonging to a Simplex to a two-dimensional Euclidean space. For a robustness check, we also tested the additive log-ratio (alr) transformation (32) described by:

$$\begin{cases} u &= \ln \frac{B\%}{H3\%} \\ v &= \ln \frac{H1\%}{H3\%}. \end{cases}$$

Alternative definition of the year: In the baseline analysis, we chose week 17 as the start of the influenza year. In the scenario *fall-year*, we take week 37 as the beginning of the year.

Alternative criterion for country/year inclusion: In the baseline analysis, we included countries/years with at least 50 positive samples. In the scenario *500+ annual cases*, we analysed countries/years with at least 500 cases.

Selection of country trajectories with highly stable classification: For the forecast, we tested an additional sensitivity scenario by limiting predictions to 50% of the countries with the most stable clustering. The HVAR model's forecasts for 2017-2019 were preceded by clustering based on country trajectories from 2010-2016, 2010-2017, and 2010-2018. For each prediction year, we calculated country-wise silhouette scores (33) to assess classification quality and selected countries that consistently exceeded a set threshold, ensuring only the most robust 50% were included.

**Alternative computations of the *mixing score* over time**

For the analysis of the mixing score, we tested its robustness both on trajectories defined in *alr* coordinates and using the Shannon entropy as an alternative metric (Fig. S8). Considering a vector of three percentages $p=(p_1,p_2,p_3)$, such that $p_1+p_2+p_3=1$, the Shannon entropy is defined as $H(p) = -\sum_{i=1}^{3} p_i \log p_i$ (34). It measures the distance of a discrete distribution from the uniform distribution. Thus, in our case, it tells us how far the relative abundances of the (sub)types are from perfect co-dominance (1/3,1/3,1/3). The distributions over time of both scores (Fig. S8, top and bottom panels, respectively) are very similar to those obtained with the isometric log-ratio transformation in Fig. 2B of the main paper.

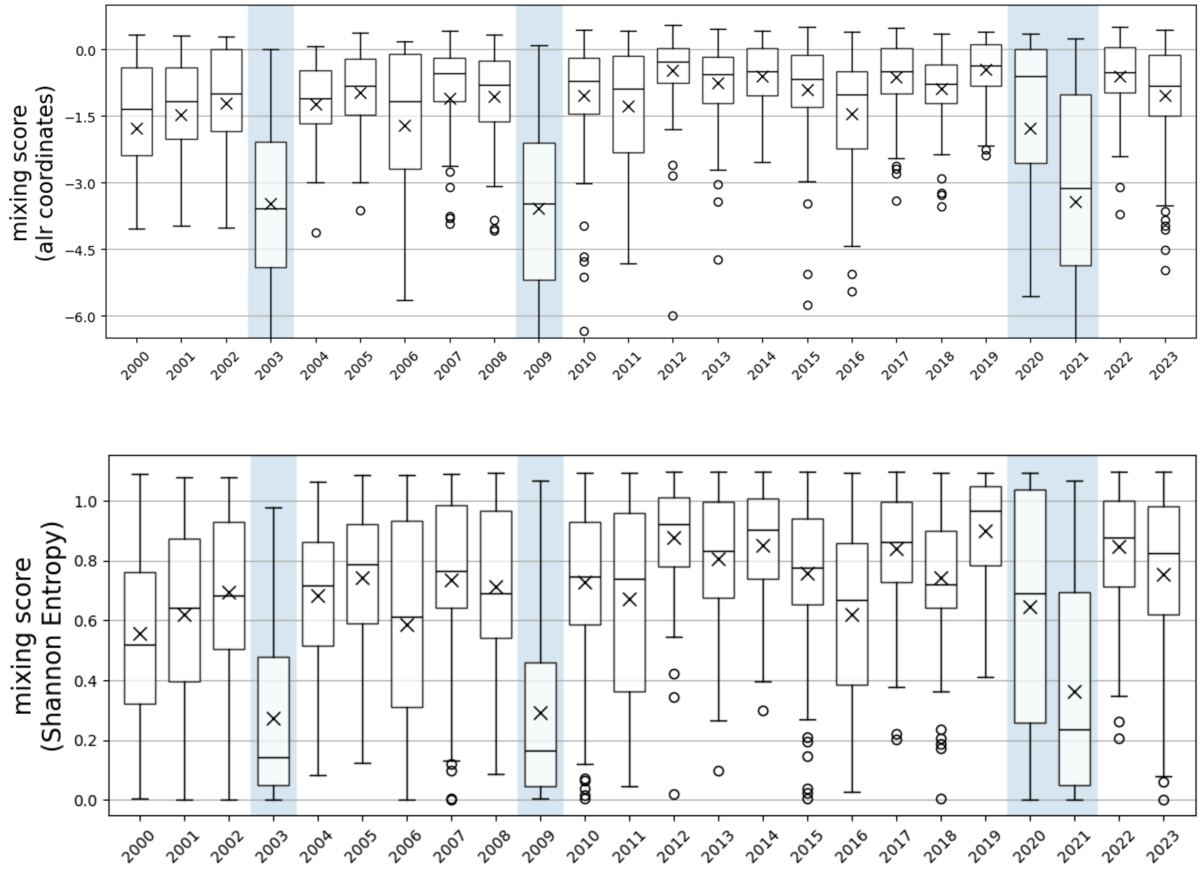


**Fig. S8 - Alternative analyses of the distributions of the degree of (sub)type mixing over time.** (Sub)type mixing is calculated via the *mixing score* under *alr* transformation (top graph), and the Shannon entropy (bottom graph).

### Geographical drivers of country similarities in (sub)type compositions under alternative scenarios

We report below (Table S4) the sensitivity analysis results regarding the association of air travel, climatic conditions, and epidemic synchrony with similarities of (sub)type trajectories. The results for the baseline scenario are compared with three alternative scenarios. The covariates are unchanged for all scenarios, while the response variable changes from one scenario to another due to different trajectory definitions.

| scenario | BASE | | 500+ annual cases | | alr coordinates | | fall-year | |
|---|---|---|---|---|---|---|---|---|
| variable | coefficient | p-value | coefficient | p-value | coefficient | p-value | coefficient | p-value |
| Intercept | -0.13 | 0.000 | -0.18 | 0.021 | -0.16 | 0.000 | 0.08 | 0.006 |
| Synchrony (semi-synchronous) | 0.14 | 0.011 | 0.10 | 0.285 | 0.19 | 0.001 | -0.18 | 0.002 |
| Synchrony (asynchronous) | 0.76 | 0.000 | 0.81 | 0.000 | 0.81 | 0.000 | 0.09 | 0.101 |
| Air-traffic distance | 0.30 | 0.000 | 0.39 | 0.000 | 0.26 | 0.000 | 0.23 | 0.000 |
| Difference in temperature | 0.24 | 0.000 | 0.01 | 0.462 | 0.24 | 0.000 | 0.43 | 0.000 |
| Difference in relative humidity | 0.10 | 0.000 | -0.03 | 0.319 | 0.11 | 0.000 | 0.03 | 0.026 |

**Table S4 - Coefficient of the multivariable regression for predicting the distance between countries' (sub)type composition trajectories in the base scenario and the alternative scenarios tested for sensitivity.**

#### Alternative trajectory clusterings

We performed the clustering for the alternative scenarios summarised in Table S3. In addition, we tested different clustering methods. Below, we refer to the classification described in the main paper as *the reference classification.*

Alternative scenarios

Below, we used the same clustering method considered in the main paper, i.e., Ward's linkage hierarchical clustering.

a. We clustered 86 country trajectories expressed in *alr* coordinates. In this scenario, the optimal partition consisted of three groups: Qatar clustered alone, and the other two groups (42 and 43 countries, respectively) were identical to the groups of the reference classification, with the exception of South Korea.

b. We clustered country trajectories of annual (sub)type composition by considering an influenza year starting in autumn (week 37). In this scenario, we defined trajectories for 88 countries that reported at least 50 cases for each year between 2010 and 2018. Of these, 84 were also included in the reference scenario, so their partitioning was comparable to the reference classification. These 84 trajectories clustered into two groups of 34 and 50 countries, which largely overlapped with the reference classification. The classifications were discordant for eight countries: Croatia, Egypt, Iran, Jordan, Kazakhstan, the Russian Federation, and Turkey.

c. We clustered trajectories for the 29 countries reporting at least 500 annual cases of flu classified by (sub)types for each year from 2010 to 2019. The optimal partition identified two groups, with only the Russian Federation changing group compared to the reference classification.

Alternative clustering methods

The clustering analysis of Fig. 3 of the main paper was obtained with the Ward's linkage algorithm. We tested two alternative methods for clustering the same 86 trajectories considered in the reference classification.

W*eighted linkage hierarchical clustering* (35): The algorithm separated Qatar and then Nicaragua from all other countries in the first two steps. The next iteration led to a decrease in the Silhouette score, resulting in a sub-optimal partition that largely coincides with the reference classification. In particular, the remaining 84 countries were divided into two groups matching the groups in the reference classification, with the exception of El Salvador.

*K-medoid clustering:* This algorithm corresponds to the better-known k-means method, where barycenters are substituted with medoids - see chp. Partitioning Around Medoids in (36). We found a 2-group partition with 36 and 50 countries in each group. This was similar to the reference classification except for seven countries that changed group: Croatia, Israel, Jordan, Mongolia, the Russian Federation, Tunisia, and Turkey.

**Alternative forecasting analyses and forecasting performance computations**

Hereafter, we provide a sensitivity analysis of the forecast of the next year's (sub)type abundances. First, we evaluated the performance of the forecasting models using different metrics for all observables. Results are shown in Table S5 ((sub)type compositions and dominance states) and Table S6 (dominance/negligibility of each (sub)type). In addition, we repeated the predictions with the trajectories defined under four alternative scenarios (Table S7).

Model ranking for dominance state and (sub)type composition was robust to the different performance metrics, with the exception of the predictions of the dominance state for Group I countries, which were better predicted by the *M3 average composition* method according to the alternative evaluation metrics (Table S5).

The best-performing model was less stable in varying evaluation methods in the case of binary predictions (Table S6). However, the key findings discussed in the main paper were robust, i.e., better predictions were obtained for A/H3N2 dominance and negligibility and B negligibility. In these cases, the *M5 HVAR* was the best-performing model in the majority of cases.

Predictions for four alternative scenarios are compared to the baseline in Table S7. The analysis confirms that:

(i) The M5 HVAR was the best-performing algorithm for most observables. For 6 out of 8 observables, M5 was the best model for almost all scenarios. Forecasting performances of H1 dominance and negligibility were, in general, lower than the other (sub)types for all scenarios, and the best performing model was less robust in varying the scenario.

(ii) H1 dominance/negligibility is harder to predict than B and H3.

Notably, predictions were better for the 50% best-classified countries than for the other scenarios, for most of the observables. For example, accuracy in predicting the dominance state improved from 34% to 38% compared to the baseline, and AUROC from 0.62 to 0.82 for B dominance. This improvement appears to be more pronounced for the M5 method than for the other methods, suggesting that selecting countries that are consistently similar over time helps to improve the spatial information used to feed the HVAR model.

| | | Performances in predicting the (sub)type composition | | | | Performances in predicting the dominance state | | |
|---|---|---|---|---|---|---|---|---|
| **group** | **method** | **ES** | **DSS** | **VS05** | **VS1** | **Accuracy** | **AUROC** | **Average precision (from the P-R curve)** |
| Group I (40) | M1 past frequencies | / | / | / | / | 0.235 ± 0.003 | 0.476 ± 0.002 | 0.325 ± 0.001 |
| | M2 H1-H3 alternation | 3.813 ± 0.011 | / | / | / | 0.227 ± 0.003 | / | / |
| | M3 avg composition | 1.811 ± 0.009 | 32.36 ± 0.36 | 0.985 ± 0.009 | 7.09 ± 0.09 | 0.218 ± 0.003 | 0.475 ± 0.002 | 0.308 ± 0.001 |
| | M4 VAR | 2.194 ± 0.009 | 8.80 ± 0.11 | 1.498 ± 0.009 | 14.52 ± 0.10 | 0.193 ± 0.003 | 0.457 ± 0.002 | 0.322 ± 0.002 |
| | M5 HVAR | **1.426 ± 0.006** | **4.42 ± 0.03** | **0.885 ± 0.006** | **6.37 ± 0.06** | **0.277 ± 0.003** | **0.577 ± 0.002** | **0.382 ± 0.002** |
| Group II (45) | M1 past frequencies | / | / | / | / | 0.301 ± 0.002 | 0.557 ± 0.002 | 0.367 ± 0.002 |
| | M2 H1-H3 alternation | 2.102 ± 0.009 | / | / | / | 0.301 ± 0.003 | / | / |
| | M3 avg composition | 1.338 ± 0.007 | 30.08 ± 0.37 | 0.739 ± 0.008 | 5.31 ± 0.09 | 0.353 ± 0.003 | **0.580 ± 0.002** | **0.407 ± 0.002** |
| | M4 VAR | 1.640 ± 0.008 | 8.52 ± 0.10 | 1.224 ± 0.013 | 13.63 ± 0.22 | 0.257 ± 0.003 | 0.514 ± 0.002 | 0.357 ± 0.002 |
| | M5 HVAR | **1.195 ± 0.006** | **3.21 ± 0.03** | **0.674 ± 0.005** | **4.95 ± 0.06** | **0.397 ± 0.003** | 0.552 ± 0.003 | 0.374 ± 0.002 |
| all countries (85) | M1 past frequencies | / | / | / | / | 0.271 ± 0.002 | 0.532 ± 0.002 | 0.345 ± 0.001 |
| | M2 H1-H3 alternation | 2.901 ± 0.008 | / | / | / | 0.267 ± 0.002 | / | / |
| | M3 avg composition | 1.559 ± 0.005 | 31.15 ± 0.25 | 0.854 ± 0.005 | 6.14 ± 0.06 | 0.290 ± 0.002 | 0.553 ± 0.002 | 0.365 ± 0.001 |
| | M4 VAR | 1.899 ± 0.006 | 8.65 ± 0.07 | 1.352 ± 0.009 | 14.05 ± 0.15 | 0.227 ± 0.002 | 0.518 ± 0.002 | 0.338 ± 0.001 |
| | M5 HVAR | **1.303 ± 0.004** | **3.77 ± 0.02** | **0.772 ± 0.004** | **5.61 ± 0.05** | **0.341 ± 0.002** | **0.613 ± 0.001** | **0.381 ± 0.001** |

**Table S5 - Robustness of model performances for predicting the (sub)type compositions and the dominance states.** Four different scores are shown to evaluate the performances in predicting the (sub)type compositions: the Energy Score considered in the manuscript and three alternative metrics. All these four metrics are negatively oriented. Similarly, the performances for predicting dominance states are presented in terms of Accuracy, the main metric used in the manuscript, and two alternative scores. These three metrics are positively oriented. The best performing methods by country grouping and metric are shown in bold. All metrics are defined in Table S1.

| group | method | Precision | AUROC | Average precision (from the P-R curve) | Precision | AUROC | Average precision (from the P-R curve) |
|---|---|---|---|---|---|---|---|
| | | **B will be dominant (>50%): yes/no ?** | | | **B will be negligible (<10%): yes/no ?** | | |
| Group I (40) | M1 past frequencies | / | 0.452 ± 0.004 | 0.268 ± 0.004 | / | 0.412 ± 0.004 | 0.357 ± 0.004 |
| | M2 H1-H3 alternation | 0.000 ± 0.000 | / | / | 0.094 ± 0.003 | / | / |
| | M3 avg composition | 0.000 ± 0.000 | 0.495 ± 0.005 | 0.236 ± 0.004 | 0.000 ± 0.000 | 0.349 ± 0.003 | 0.278 ± 0.003 |
| | M4 VAR | 0.150 ± 0.006 | 0.425 ± 0.004 | 0.203 ± 0.003 | 0.500 ± 0.008 | 0.610 ± 0.004 | 0.436 ± 0.005 |
| | M5 HVAR | **0.217 ± 0.007** | **0.641 ± 0.003** | **0.288 ± 0.004** | **0.800 ± 0.007** | **0.905 ± 0.002** | **0.849 ± 0.003** |
| Group II (45) | M1 past frequencies | / | **0.715 ± 0.006** | **0.125 ± 0.004** | / | 0.521 ± 0.006 | 0.173 ± 0.004 |
| | M2 H1-H3 alternation | 0.000 ± 0.000 | / | / | 0.000 ± 0.000 | / | / |
| | M3 avg composition | 0.000 ± 0.000 | 0.693 ± 0.005 | 0.118 ± 0.004 | 0.125 ± 0.009 | 0.507 ± 0.005 | 0.154 ± 0.004 |
| | M4 VAR | **0.053 ± 0.004** | 0.463 ± 0.008 | 0.084 ± 0.004 | **0.143 ± 0.005** | 0.527 ± 0.004 | 0.145 ± 0.003 |
| | M5 HVAR | 0.000 ± 0.000 | 0.368 ± 0.009 | 0.065 ± 0.002 | 0.000 ± 0.000 | **0.648 ± 0.004** | **0.185 ± 0.003** |
| all countries (85) | M1 past frequencies | / | 0.551 ± 0.004 | 0.183 ± 0.003 | / | 0.561 ± 0.003 | 0.289 ± 0.003 |
| | M2 H1-H3 alternation | 0.000 ± 0.000 | / | / | 0.073 ± 0.002 | / | / |
| | M3 avg composition | 0.000 ± 0.000 | **0.624 ± 0.003** | 0.183 ± 0.002 | 0.111 ± 0.011 | 0.506 ± 0.003 | 0.225 ± 0.002 |
| | M4 VAR | 0.103 ± 0.003 | 0.496 ± 0.003 | 0.142 ± 0.002 | 0.300 ± 0.004 | 0.603 ± 0.003 | 0.277 ± 0.002 |
| | M5 HVAR | **0.208 ± 0.006** | 0.618 ± 0.003 | **0.212 ± 0.003** | **0.522 ± 0.007** | **0.815 ± 0.002** | **0.573 ± 0.005** |
| | | **H1 will be dominant (>50%): yes/no ?** | | | **H1 will be negligible (<10%): yes/no ?** | | |
| Group I (40) | M1 past frequencies | / | **0.489 ± 0.004** | **0.398 ± 0.004** | / | **0.699 ± 0.006** | **0.206 ± 0.006** |
| | M2 H1-H3 alternation | 0.263 ± 0.004 | / | / | **0.053 ± 0.004** | / | / |
| | M3 avg composition | **1.000 ± 0.034** | 0.414 ± 0.004 | 0.353 ± 0.005 | 0.000 ± 0.000 | 0.645 ± 0.006 | 0.167 ± 0.005 |
| | M4 VAR | 0.097 ± 0.004 | 0.327 ± 0.003 | 0.281 ± 0.003 | 0.023 ± 0.002 | 0.281 ± 0.005 | 0.072 ± 0.002 |
| | M5 HVAR | 0.316 ± 0.008 | 0.480 ± 0.004 | 0.341 ± 0.004 | 0.053 ± 0.003 | 0.253 ± 0.006 | 0.070 ± 0.002 |
| Group II (45) | M1 past frequencies | / | 0.440 ± 0.004 | 0.354 ± 0.004 | / | 0.588 ± 0.003 | 0.188 ± 0.003 |
| | M2 H1-H3 alternation | **0.415 ± 0.005** | / | / | 0.267 ± 0.009 | / | / |
| | M3 avg composition | 0.364 ± 0.010 | 0.505 ± 0.004 | 0.364 ± 0.004 | 0.286 ± 0.013 | 0.580 ± 0.004 | 0.196 ± 0.005 |
| | M4 VAR | 0.300 ± 0.006 | 0.527 ± 0.004 | 0.351 ± 0.004 | 0.143 ± 0.004 | 0.548 ± 0.005 | 0.162 ± 0.003 |
| | M5 HVAR | 0.300 ± 0.011 | **0.584 ± 0.004** | **0.408 ± 0.005** | **0.455 ± 0.012** | **0.605 ± 0.006** | **0.278 ± 0.007** |
| all countries (85) | M1 past frequencies | / | 0.481 ± 0.003 | **0.366 ± 0.003** | / | **0.596 ± 0.004** | **0.169 ± 0.003** |
| | M2 H1-H3 alternation | 0.327 ± 0.003 | / | / | 0.147 ± 0.004 | / | / |
| | M3 avg composition | **0.417 ± 0.010** | 0.485 ± 0.003 | 0.346 ± 0.003 | **0.250 ± 0.012** | 0.567 ± 0.004 | 0.160 ± 0.004 |
| | M4 VAR | 0.197 ± 0.004 | 0.439 ± 0.003 | 0.300 ± 0.002 | 0.076 ± 0.002 | 0.423 ± 0.004 | 0.105 ± 0.001 |
| | M5 HVAR | 0.310 ± 0.006 | **0.529 ± 0.003** | 0.357 ± 0.003 | 0.200 ± 0.005 | 0.460 ± 0.004 | 0.138 ± 0.003 |
| | | **H3 will be dominant (>50%): yes/no ?** | | | **H3 will be negligible (<10%): yes/no ?** | | |
| Group I (40) | M1 past frequencies | / | 0.433 ± 0.004 | 0.216 ± 0.003 | / | 0.375 ± 0.004 | 0.215 ± 0.003 |
| | M2 H1-H3 alternation | 0.400 ± 0.007 | / | / | 0.420 ± 0.005 | / | / |
| | M3 avg composition | 0.192 ± 0.006 | 0.478 ± 0.004 | 0.228 ± 0.004 | 0.000 ± 0.000 | 0.447 ± 0.005 | 0.238 ± 0.004 |
| | M4 VAR | 0.318 ± 0.005 | 0.640 ± 0.003 | 0.342 ± 0.005 | 0.405 ± 0.006 | 0.715 ± 0.003 | 0.375 ± 0.005 |
| | M5 HVAR | **0.472 ± 0.006** | **0.764 ± 0.003** | **0.402 ± 0.005** | **0.467 ± 0.009** | **0.796 ± 0.003** | **0.518 ± 0.006** |
| Group II (45) | M1 past frequencies | / | 0.552 ± 0.004 | 0.265 ± 0.005 | / | 0.620 ± 0.004 | 0.170 ± 0.003 |
| | M2 H1-H3 alternation | 0.318 ± 0.005 | / | / | 0.233 ± 0.006 | / | / |
| | M3 avg composition | 0.279 ± 0.005 | 0.576 ± 0.004 | 0.253 ± 0.003 | 0.000 ± 0.000 | 0.553 ± 0.005 | 0.169 ± 0.005 |
| | M4 VAR | 0.239 ± 0.004 | 0.511 ± 0.004 | 0.240 ± 0.003 | 0.182 ± 0.008 | 0.654 ± 0.006 | 0.222 ± 0.005 |
| | M5 HVAR | **0.354 ± 0.005** | **0.618 ± 0.005** | **0.345 ± 0.005** | **0.333 ± 0.022** | **0.655 ± 0.005** | **0.286 ± 0.007** |
| all countries (85) | M1 past frequencies | / | 0.506 ± 0.003 | 0.231 ± 0.003 | / | 0.565 ± 0.003 | 0.202 ± 0.002 |
| | M2 H1-H3 alternation | 0.351 ± 0.004 | / | / | 0.350 ± 0.004 | / | / |
| | M3 avg composition | 0.246 ± 0.004 | 0.539 ± 0.003 | 0.240 ± 0.002 | 0.000 ± 0.000 | 0.587 ± 0.003 | 0.220 ± 0.003 |
| | M4 VAR | 0.278 ± 0.003 | 0.578 ± 0.003 | 0.268 ± 0.003 | 0.354 ± 0.005 | 0.712 ± 0.003 | 0.309 ± 0.004 |
| | M5 HVAR | **0.405 ± 0.004** | **0.702 ± 0.002** | **0.366 ± 0.004** | **0.444 ± 0.009** | **0.758 ± 0.003** | **0.438 ± 0.005** |

**Table S6 - Robustness of models' performance for predicting the dominance (yes/no) and the negligibility (yes/no) of each subtype in varying performance evaluation methods.** The two alternative scores tested are compared with the AUROC reported in the main paper. All three metrics are positively oriented. The best performing methods by country grouping and metric are shown in bold.

| scenario | observable | metric | M1 past frequencies | M2 H1-H3 alternation | M3 avg composition | M4 VAR | M5 HVAR |
|---|---|---|---|---|---|---|---|
| BASE | composition | ES | / | 2.901 ± 0.008 | 1.559 ± 0.005 | 1.899 ± 0.006 | **1.303 ± 0.004** |
| SENSITIVITY 500+ | composition | ES | / | 2.657 ± 0.011 | 1.412 ± 0.008 | 1.653 ± 0.007 | **1.136 ± 0.004** |
| SENSITIVITY alr coordinates | composition | ES | / | 3.322 ± 0.009 | 2.135 ± 0.008 | 2.495 ± 0.008 | **1.687 ± 0.005** |
| SENSITIVITY fall-year | composition | ES | / | 3.932 ± 0.017 | 1.759 ± 0.007 | 2.137 ± 0.010 | **1.409 ± 0.005** |
| SENSITIVITY best classified | composition | ES | / | 3.051 ± 0.011 | 1.559 ± 0.009 | 1.742 ± 0.006 | **1.157 ± 0.005** |
| BASE | dominance state | Accuracy | 0.271 ± 0.002 | 0.267 ± 0.002 | 0.290 ± 0.002 | 0.227 ± 0.002 | **0.341 ± 0.002** |
| SENSITIVITY 500+ | dominance state | Accuracy | 0.322 ± 0.004 | 0.276 ± 0.004 | **0.368 ± 0.004** | 0.241 ± 0.004 | 0.333 ± 0.004 |
| SENSITIVITY alr coordinates | dominance state | Accuracy | 0.271 ± 0.002 | 0.264 ± 0.002 | 0.291 ± 0.002 | 0.229 ± 0.002 | **0.360 ± 0.002** |
| SENSITIVITY fall-year | dominance state | Accuracy | 0.293 ± 0.002 | 0.253 ± 0.002 | 0.195 ± 0.002 | 0.328 ± 0.002 | **0.339 ± 0.002** |
| SENSITIVITY best classified | dominance state | Accuracy | 0.295 ± 0.003 | 0.264 ± 0.003 | 0.349 ± 0.003 | 0.248 ± 0.003 | **0.380 ± 0.003** |
| BASE | B dominance | AUROC | 0.551 ± 0.003 | / | **0.624 ± 0.003** | 0.496 ± 0.004 | 0.618 ± 0.004 |
| SENSITIVITY 500+ | B dominance | AUROC | 0.603 ± 0.007 | / | 0.706 ± 0.006 | 0.478 ± 0.007 | **0.734 ± 0.005** |
| SENSITIVITY alr coordinates | B dominance | AUROC | 0.555 ± 0.003 | / | **0.635 ± 0.003** | 0.503 ± 0.004 | 0.571 ± 0.004 |
| SENSITIVITY fall-year | B dominance | AUROC | 0.590 ± 0.003 | / | 0.602 ± 0.004 | 0.642 ± 0.004 | **0.762 ± 0.004** |
| SENSITIVITY best classified | B dominance | AUROC | 0.539 ± 0.005 | / | 0.632 ± 0.004 | 0.668 ± 0.004 | **0.824 ± 0.004** |
| BASE | B negligibility | AUROC | 0.561 ± 0.003 | / | 0.506 ± 0.003 | 0.603 ± 0.003 | **0.815 ± 0.002** |
| SENSITIVITY 500+ | B negligibility | AUROC | 0.530 ± 0.005 | / | 0.461 ± 0.005 | 0.685 ± 0.005 | **0.813 ± 0.005** |
| SENSITIVITY alr coordinates | B negligibility | AUROC | 0.566 ± 0.003 | / | 0.513 ± 0.003 | 0.611 ± 0.003 | **0.794 ± 0.002** |
| SENSITIVITY fall-year | B negligibility | AUROC | 0.622 ± 0.003 | / | 0.630 ± 0.003 | 0.661 ± 0.003 | **0.845 ± 0.002** |
| SENSITIVITY best classified | B negligibility | AUROC | 0.564 ± 0.004 | / | 0.486 ± 0.004 | 0.683 ± 0.004 | **0.934 ± 0.002** |
| BASE | H1 dominance | AUROC | 0.481 ± 0.003 | / | 0.485 ± 0.003 | 0.439 ± 0.003 | **0.529 ± 0.003** |
| SENSITIVITY 500+ | H1 dominance | AUROC | 0.454 ± 0.005 | / | 0.473 ± 0.005 | 0.440 ± 0.004 | **0.526 ± 0.004** |
| SENSITIVITY alr coordinates | H1 dominance | AUROC | 0.481 ± 0.003 | / | 0.482 ± 0.003 | 0.435 ± 0.002 | **0.572 ± 0.002** |
| SENSITIVITY fall-year | H1 dominance | AUROC | 0.530 ± 0.003 | / | **0.534 ± 0.003** | 0.446 ± 0.003 | 0.391 ± 0.003 |
| SENSITIVITY best classified | H1 dominance | AUROC | 0.407 ± 0.004 | / | 0.423 ± 0.004 | 0.377 ± 0.003 | **0.555 ± 0.004** |
| BASE | H1 negligibility | AUROC | **0.596 ± 0.003** | / | 0.567 ± 0.004 | 0.423 ± 0.004 | 0.460 ± 0.005 |
| SENSITIVITY 500+ | H1 negligibility | AUROC | 0.517 ± 0.006 | / | **0.600 ± 0.005** | 0.329 ± 0.005 | 0.411 ± 0.008 |
| SENSITIVITY alr coordinates | H1 negligibility | AUROC | **0.601 ± 0.003** | / | 0.571 ± 0.004 | 0.433 ± 0.003 | 0.477 ± 0.004 |
| SENSITIVITY fall-year | H1 negligibility | AUROC | **0.588 ± 0.005** | / | 0.570 ± 0.005 | 0.482 ± 0.006 | 0.394 ± 0.006 |
| SENSITIVITY best classified | H1 negligibility | AUROC | **0.673 ± 0.005** | / | 0.600 ± 0.006 | 0.459 ± 0.005 | 0.463 ± 0.007 |
| BASE | H3 dominance | AUROC | 0.506 ± 0.003 | / | 0.539 ± 0.003 | 0.578 ± 0.003 | **0.702 ± 0.003** |
| SENSITIVITY 500+ | H3 dominance | AUROC | 0.542 ± 0.005 | / | 0.589 ± 0.005 | 0.634 ± 0.005 | **0.724 ± 0.005** |
| SENSITIVITY alr coordinates | H3 dominance | AUROC | 0.513 ± 0.003 | / | 0.543 ± 0.003 | 0.580 ± 0.003 | **0.713 ± 0.003** |
| SENSITIVITY fall-year | H3 dominance | AUROC | 0.506 ± 0.003 | / | 0.491 ± 0.004 | 0.669 ± 0.003 | **0.717 ± 0.003** |
| SENSITIVITY best classified | H3 dominance | AUROC | 0.535 ± 0.004 | / | 0.576 ± 0.004 | 0.579 ± 0.004 | **0.685 ± 0.004** |
| BASE | H3 negligibility | AUROC | 0.565 ± 0.003 | / | 0.587 ± 0.003 | 0.712 ± 0.003 | **0.758 ± 0.003** |
| SENSITIVITY 500+ | H3 negligibility | AUROC | 0.573 ± 0.005 | / | 0.597 ± 0.006 | 0.754 ± 0.005 | **0.799 ± 0.005** |
| SENSITIVITY alr coordinates | H3 negligibility | AUROC | 0.557 ± 0.003 | / | 0.566 ± 0.003 | 0.702 ± 0.003 | **0.740 ± 0.003** |
| SENSITIVITY fall-year | H3 negligibility | AUROC | 0.511 ± 0.004 | / | 0.528 ± 0.004 | 0.728 ± 0.003 | **0.735 ± 0.003** |
| SENSITIVITY best classified | H3 negligibility | AUROC | 0.573 ± 0.004 | / | 0.581 ± 0.004 | 0.733 ± 0.004 | **0.762 ± 0.004** |

**Table S7 - Robustness of models' performances in providing predictions for alternative scenarios.** Performances for the base scenario are compared with those for four alternative scenarios defined in Table S3. For each scenario, the forecasting methods are evaluated on the predictions made for eight observables: the (sub)type composition, the subtypes, the dominance state, and the dominance and negligibility of each subtype. For each observable, the same metric considered in the manuscript was used to assess the goodness of the prediction. Methods performing best by scenario are in bold.

### Additional Bibliography


1. J. Tamerius, *et al.*, Global Influenza Seasonality: Reconciling Patterns across Temperate and Tropical Regions. *Environ. Health Perspect.* **119**, 439–445 (2010).

2. F. Bonacina, *et al.*, Global patterns and drivers of influenza decline during the COVID-19 pandemic. *Int. J. Infect. Dis.* **128**, 132–139 (2023).

3. J.-A. Martín-Fernández, K. Hron, M. Templ, P. Filzmoser, J. Palarea-Albaladejo, Bayesian-multiplicative treatment of count zeros in compositional data sets. *Stat.*

*Model.* **15**, 134–158 (2015).

4. J. Palarea-Albaladejo, J. A. Martín-Fernández, zCompositions — R package for multivariate imputation of left-censored data under a compositional approach. *Chemom. Intell. Lab. Syst.* **143**, 85–96 (2015).

5. M. Del Riccio, *et al.*, Global analysis of respiratory viral circulation and timing of epidemics in the pre–COVID-19 and COVID-19 pandemic eras, based on data from the Global Influenza Surveillance and Response System (GISRS). *Int. J. Infect. Dis.* **144**, 107052 (2024).

6. G. G. Koch, "Intraclass Correlation Coefficient" in *Encyclopedia of Statistical Sciences*, (John Wiley & Sons, Ltd, 2004).

7. A. Gautreau, A. Barrat, M. Barthélemy, Global disease spread: Statistics and estimation of arrival times. *J. Theor. Biol.* **251**, 509–522 (2008).

8. B. Faucher, *et al.*, Drivers and impact of the early silent invasion of SARS-CoV-2 Alpha. *Nat. Commun.* **15**, 2152 (2024).

9. United Nations, Department of Economic and Social Affairs, Population Division, World Population Prospects 2024, Online Edition. (2024). Available at: https://population.un.org/wpp/downloads?folder=Standard%20Projections&group=Most%20used [Accessed 27 March 2025].

10. D. Brockmann, D. Helbing, The Hidden Geometry of Complex, Network-Driven Contagion Phenomena. *Science* **342**, 1337–1342 (2013).

11. H. Hersbach, *et al.*, ERA5 monthly averaged data on pressure levels from 1940 to present. https://doi.org/10.24381/cds.6860a573.

12. United Nations, Department of Economic and Social Affairs, Population Division, World Urbanization Prospects: The 2018 Revision. File 12: Population of Urban Agglomerations with 300,000 Inhabitants or More in 2018, by Country, 1950-2035 (thousands). (2018). Available at: https://population.un.org/wup/Download/ [Accessed 30 April 2022].

13. United Nations, Department of Economic and Social Affairs, Population Division, World Urbanization Prospects: The 2018 Revision. File 13: Population of Capital Cities in 2018 (thousands). (2018). Available at: https://population.un.org/wup/Download/ [Accessed 30 April 2022].

14. N. Mantel, The detection of disease clustering and a generalized regression approach. *Cancer Res.* (1967).

15. J. W. Lichstein, Multiple regression on distance matrices: a multivariate spatial analysis tool. *Plant Ecol.* **188**, 117–131 (2007).

16. H. Lütkepohl, *New introduction to multiple time series analysis: with ... 36 tables*, 1. ed., corr. 2. print (Springer, 2007).

17. F. Lu, Y. Zheng, H. Cleveland, C. Burton, D. Madigan, Bayesian hierarchical vector autoregressive models for patient-level predictive modeling. *PLOS ONE* **13**, e0208082 (2018).

18. T. Gneiting, A. E. Raftery, Strictly Proper Scoring Rules, Prediction, and Estimation. *J. Am. Stat. Assoc.* **102**, 359–378 (2007).

19. J. Bracher, E. L. Ray, T. Gneiting, N. G. Reich, Evaluating epidemic forecasts in an interval format. *PLOS Comput. Biol.* **17**, e1008618 (2021).

20. M. Scheuerer, T. M. Hamill, Variogram-Based Proper Scoring Rules for Probabilistic Forecasts of Multivariate Quantities. *Mon. Weather Rev.* **143**, 1321–1334 (2015).

21. F. Pedregosa, *et al.*, Scikit-learn: Machine Learning in Python. *J. Mach. Learn. Res.* **12**, 2825–2830 (2011).

22. V. Dhanasekaran, *et al.*, Human seasonal influenza under COVID-19 and the potential consequences of influenza lineage elimination. *Nat. Commun.* **13**, 1721 (2022).

23. P. Zanobini, *et al.*, Global patterns of seasonal influenza activity, duration of activity and virus (sub)type circulation from 2010 to 2020. *Influenza Other Respir. Viruses* **16**, 696–706 (2022).

24. L. Zheng, *et al.*, Global variability of influenza activity and virus subtype circulation from 2011 to 2023. *BMJ Open Respir. Res.* **10**, e001638 (2023).

25. T. Bedford, *et al.*, Global circulation patterns of seasonal influenza viruses vary with antigenic drift. *Nature* **523**, 217–220 (2015).

26. V. N. Petrova, C. A. Russell, The evolution of seasonal influenza viruses. *Nat. Rev. Microbiol.* **16**, 47–60 (2018).

27. W. Yang, E. H. Y. Lau, B. J. Cowling, Dynamic interactions of influenza viruses in Hong Kong during 1998-2018. *PLOS Comput. Biol.* **16**, e1007989 (2020).

28. K. L. Laurie, *et al.*, Interval Between Infections and Viral Hierarchy Are Determinants of Viral Interference Following Influenza Virus Infection in a Ferret Model. *J. Infect. Dis.* **212**, 1701–1710 (2015).

29. X.-S. Zhang, D. De Angelis, Construction of the influenza A virus transmission tree in a college-based population: co-transmission and interactions between influenza A viruses. *BMC Infect. Dis.* **16**, 38 (2016).

30. L. Gatti, *et al.*, Cross-reactive immunity potentially drives global oscillation and opposed alternation patterns of seasonal influenza A viruses. *Sci. Rep.* **12**, 8883 (2022).

31. E. Goldstein, S. Cobey, S. Takahashi, J. C. Miller, M. Lipsitch, Predicting the Epidemic Sizes of Influenza A/H1N1, A/H3N2, and B: A Statistical Method. *PLOS Med.* **8**, e1001051 (2011).

32. J. Aitchison, *The statistical analysis of compositional data* (Chapman & Hall, Ltd., 1986).

33. P. J. Rousseeuw, Silhouettes: A graphical aid to the interpretation and validation of cluster analysis. *J. Comput. Appl. Math.* **20**, 53–65 (1987).

34. C. E. Shannon, A mathematical theory of communication. *Bell Syst. Tech. J.* **27**, 623–656 (1948).

35. scipy.cluster.hierarchy.weighted — SciPy v1.13.0 Manual.

36. L. Kaufman, P. J. Rousseeuw, “Partitioning Around Medoids (Program PAM)” in *Finding Groups in Data*, (John Wiley & Sons, Ltd, 1990), pp. 68–125.
